# Accurate, sensitive, and efficient chromatin accessibility quantification at target loci using UNIChro-seq

**DOI:** 10.1101/2025.07.29.25332340

**Authors:** Michihiro Kono, Hiroaki Hatano, Kenichiro Asahara, Masahiro Nakano, Reza Bagherzadeh, Tsugumi Kawashima, Takahiro Arakawa, Miho Sato, Hajime Inokuchi, Takahiro Nishino, Takahiro Itamiya, Haruka Takahashi, Bunki Natsumoto, Akari Suzuki, Kazuhiko Yamamoto, Kazuyoshi Ishigaki

## Abstract

Recent progress in statistical and experimental fine mapping of disease risk variants prompts us to focus on specific target loci for functional investigation. However, current genetics is hindered by a limited toolbox for target-loci analysis. To address this, we developed UNIChro-seq, a method that digitally counts accessible chromatin molecules at target loci. UNIChro-seq allows for accurate, sensitive, and efficient quantification of allelic effects compared to conventional methods. Using UNIChro-seq, we investigated the effects of 57 autoimmunity risk alleles on chromatin accessibility and estimated the causal effects of 20 artificial variants generated through genome editing. As a caveat, non-negligible fraction of the edited allele exhibited a falsely positive effect on chromatin accessibility, which can be effectively distinguished from the true causal effect through bi-directional genome editing. Finally, functional dissection of a fine-mapped risk variant at the *LEF1* locus illuminated its impact on T cell pathology in rheumatoid arthritis. Together, these findings underscore the utility of combining UNIChro-seq with genome editing technology to enable precise and scalable functional analysis of disease-associated loci.

## Introduction

Genome-wide association studies (GWAS) have successfully identified numerous risk variants associated with complex diseases^1–3^. To better understand disease etiology, it is crucial to precisely clarify the molecular functions of these risk variants. Most risk variants are found in open chromatin regions of disease-relevant cell types near disease-relevant genes, suggesting that they likely contribute to disease development by altering the surrounding chromatin status and influencing gene expression. Consequently, previous studies have intensely investigated the association between risk variants and molecular phenotypes, such as chromatin accessibility and gene expression^4,5^. The most common approach is quantitative trait loci (QTL) analysis, which typically requires several hundred donors. Although many large-scale QTL studies have been conducted, including Immgen^6^, eQTLGen^7^, GTEx^8,9^, DICE^10,11^, and ImmuNexUT^12^, many risk variants still lack identified QTL effects^13,14^, referred to as “missing QTL”. These missing QTLs represent significant hurdles that hinder understanding disease pathogenesis. Given that these issues remain largely unsolved despite previous large-scale QTL studies, the functional genetics research community faces an urgent need to consider reforming the structure of the experimental systems.

Several limitations in the experimental system for quantifying molecular phenotypes may contribute to the missing QTLs. The first and most significant limitation is inefficiency; causal QTL effects may be highly cell-state specific, and conventional QTL experimental systems are not efficient enough to screen a variety of cell states. The second limitation is low sensitivity; causal molecular phenotypes could be rare events that are challenging to quantify with current experimental systems (e.g., lowly expressed genes). The third limitation is insufficient accuracy; the causal QTL effects might be very subtle, typical of common variants that have survived selective pressure^15^. These limitations could be addressed through three major shifts in experimental design (**Figure 1a**). The first shift is from population-scale to allele-specific analysis^16,17^, requiring only a few donors. Unlike population-scale analyses that require hundreds of donors, allele-specific analysis only needs a few donors, allowing for better allocation of experimental resources towards expanding cell types and facilitating time-series assessments^18^. Although conventional allele-specific analyses often suffer from noise due to donor-specific haplotype structures or linkage disequilibrium (LD), these limitations can be alleviated by incorporating genome editing technology^19,20^. The second shift involves transitioning from genome-wide to targeted-loci amplification. Targeted-loci amplification can preserve the complexity of the molecular phenotypes of interest, theoretically enhancing the sensitivity of detecting QTL effects. Additionally, targeted-loci analysis improves experimental efficiency, as typical genetics research—especially fine-mapping and genome-editing studies—focuses on the allelic effects of a limited number of variants rather than genome-wide variants. The final shift is moving from sequencing read-based to tag-based quantification, enabling digital counting. Molecular tags, such as unique molecular identifiers (UMIs), can suppress amplification bias and improve the accuracy of QTL effect estimation, which is particularly important in targeted-locus amplification protocols that typically involve multiple rounds of amplification.

**Figure 1.**
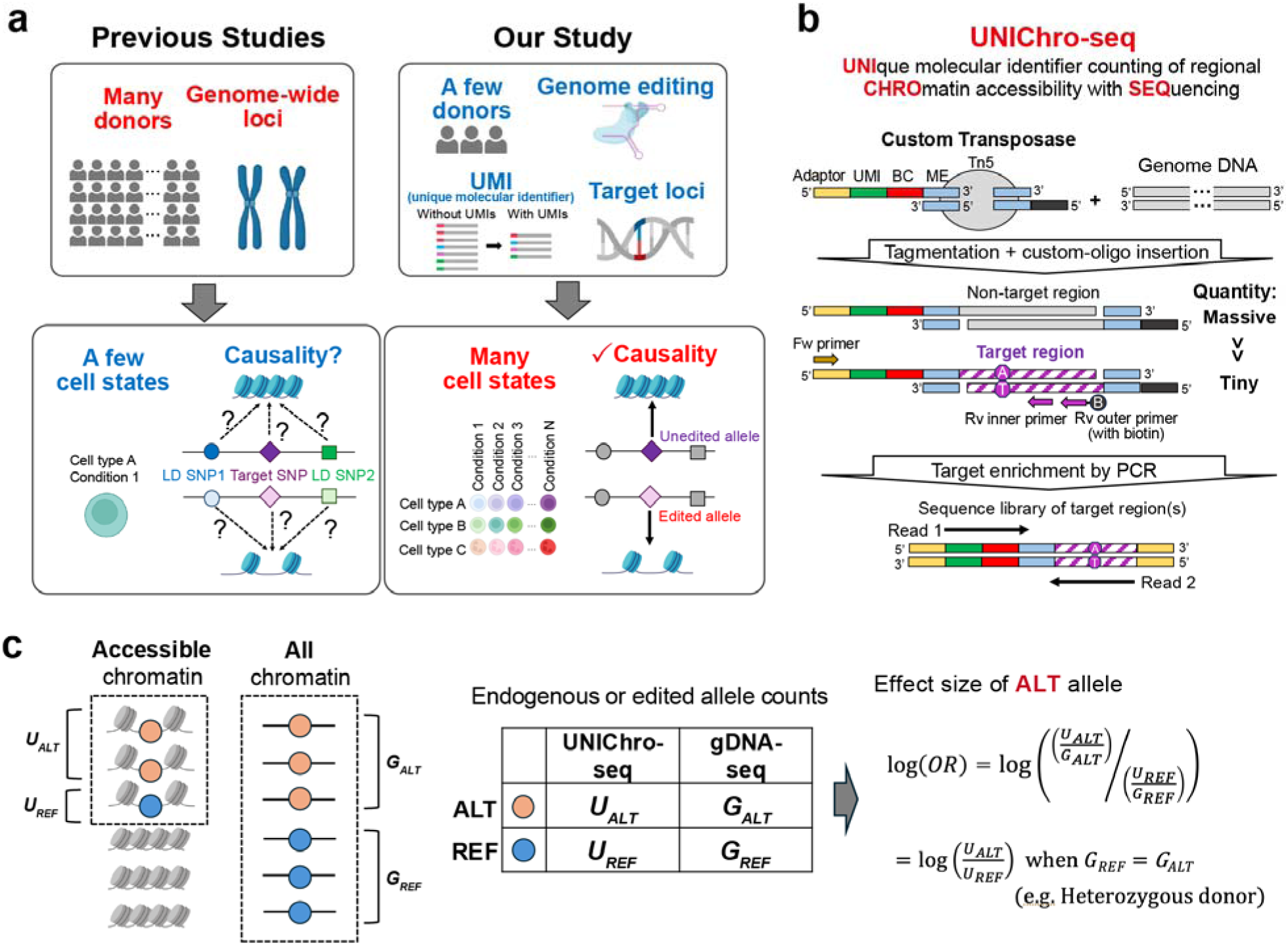
Overview of our study design and UNIChro-seq. **a**, We propose a shift from conventional genome-wide analyses across many donors to targeted locus-specific analyses in fewer donors, leveraging CRISPR genome editing and allele-specific analysis. While traditional approaches are limited by linkage disequilibrium (LD) structure and have constraints in confirming causality, our proposed strategy overcomes these limitations. Furthermore, the efficiency of allele-specific analysis enables exploration across a broader range of cell states, expanding the search space for functional insights. This panel was created with Biorender.com. **b**, We developed UNIChro-seq, which selectively amplifies specific accessible chromatin regions, accurately quantifies allelic imbalance using unique molecular indexes (UMIs), and allows us to pool multiple libraries using transposase with custom barcode (BC) adaptors. ME, mosaic end; Fw, forward; Rv, reverse. See **Supplementary Figures 1 and 2** for detailed illustration. **c**, The basic strategy for estimating allelic effects using UNIChro-seq. UNIChro-seq yields UMI-corrected values (*U_ALT_* and *U_REF_*) representing chromatin accessibility, while genomic DNA sequencing (gDNA-seq) provides read counts (*G_ALT_* and *G_REF_*) representing editing efficiency (when using endogenous heterozygous alleles, we assumed *G_ALT_* = *G_REF_*). The effect size on the chromatin accessibility of the alternative allele (ALT) compared with the reference allele (REF) is defined as the natural logarithm of the odds ratio, which can be intuitively understood from a 2 × 2 table using these values.

In this study, we aimed to address the issues related to missing QTL effects and developed an experimental platform—**UNI**que molecular identifier counting of regional **CHRO**matin accessibility with **SEQ**uencing (UNIChro-seq)—to quantify the allelic effects on chromatin accessibility at target loci (**Figure 1b**). We demonstrated that UNIChro-seq can digitally quantify molecular abundance and provide significantly more accurate, sensitive, and efficient quantification of allelic effects at target loci compared to conventional assay for transposase-accessible chromatin with high-throughput sequencing (ATAC-seq), which analyzes genome-wide chromatin accessibility. By incorporating artificial variants generated through genome editing, we experimentally fine-mapped causal variants and identified pitfalls within genome-editing experiments and the importance of bi-directional genome editing. Leveraging the efficiency of UNIChro-seq, we evaluated the allelic effects of autoimmunity risk variants through time-series experiments. A notable example involved the risk allele at the *LEF1* locus, where we introduced both risk and protective alleles bi-directionally in human CD4^+^ T cells and assessed their allelic effects under multiple cell conditions using UNIChro-seq. Additionally, we tested the immunological relevance of *LEF1* through intervention experiments utilizing single-cell analysis. Overall, our study successfully evaluated the molecular functions of 75 variants (20 of which were incorporated via genome editing) in 57 cell types and conditions, utilizing only 20 donors. This research provides valuable insights into the causal mechanisms underlying autoimmune diseases and demonstrates the feasibility and promise of a targeted approach using UNIChro-seq combined with genome editing technology.

## Results

### Establishment and benchmarking of UNIChro-seq

To accurately, sensitively, and efficiently evaluate allelic effects on the chromatin accessibility at target regions, we developed UNIChro-seq by modifying conventional ATAC-seq^21^ **(Figure 1b, Supplementary Figure 1a-c, Methods)**. Briefly, we custom-designed DNA oligonucleotides attached to Tn5 transposase, incorporating UMIs for accurate quantification and barcodes to pool multiple samples into a single sequencing library. After the customized transposase digests and tags open chromatin regions with UMIs and barcodes, the system amplifies the target regions by PCR using nested primers. To prevent unintended UMI overwriting due to detached transposase adapters and PCR chimeras during library preparation, we divide the library amplification procedure into three steps utilizing blocking primers (**Supplementary Figure 2a**). In silico, we remove PCR duplicates based on UMIs that have passed multiple quality check criteria (**Supplementary Figure 2b**), which enables the suppression of PCR bias and digital counting of accessible chromatin molecules carrying each allele. In this manuscript, we assumed that the UNIChro-seq allelic count follows a binomial distribution and applied logistic regression to estimate the effect size (**Figure 1c**). The effect allele is the alternative (ALT) allele unless stated otherwise.

The accuracy of UNIChro-seq primarily depends on UMI performance. To assess this, we first benchmarked UMI behavior in an artificial system simulating the amplification process following transposase tagmentation (**Supplementary Figure 1a**). In this system, we synthesized DNA oligonucleotides mimicking the tagmented DNA fragments at the rs2061831 locus, with equal copies of T- and C-alleles, ranging from 10 to 10^4^ copies per reaction. We compared the performance of UNIChro-seq against digital PCR, the gold standard for digital counting. The UMI counts estimated by UNIChro-seq were nearly identical to the copy numbers estimated by digital PCR (**Figure 2a**). Furthermore, the C-allele frequency quantified by UNIChro-seq consistently aligned with the expected value, with deviations falling within the theoretically expected range (**Supplementary Figure 3a, Supplementary Table 1, Methods**). Our data demonstrated that UNIChro-seq can amplify fragments with sufficient accuracy while suppressing amplification bias through UMIs, even when as few as 10 copies of alleles are present in the reaction.

**Figure 2.**
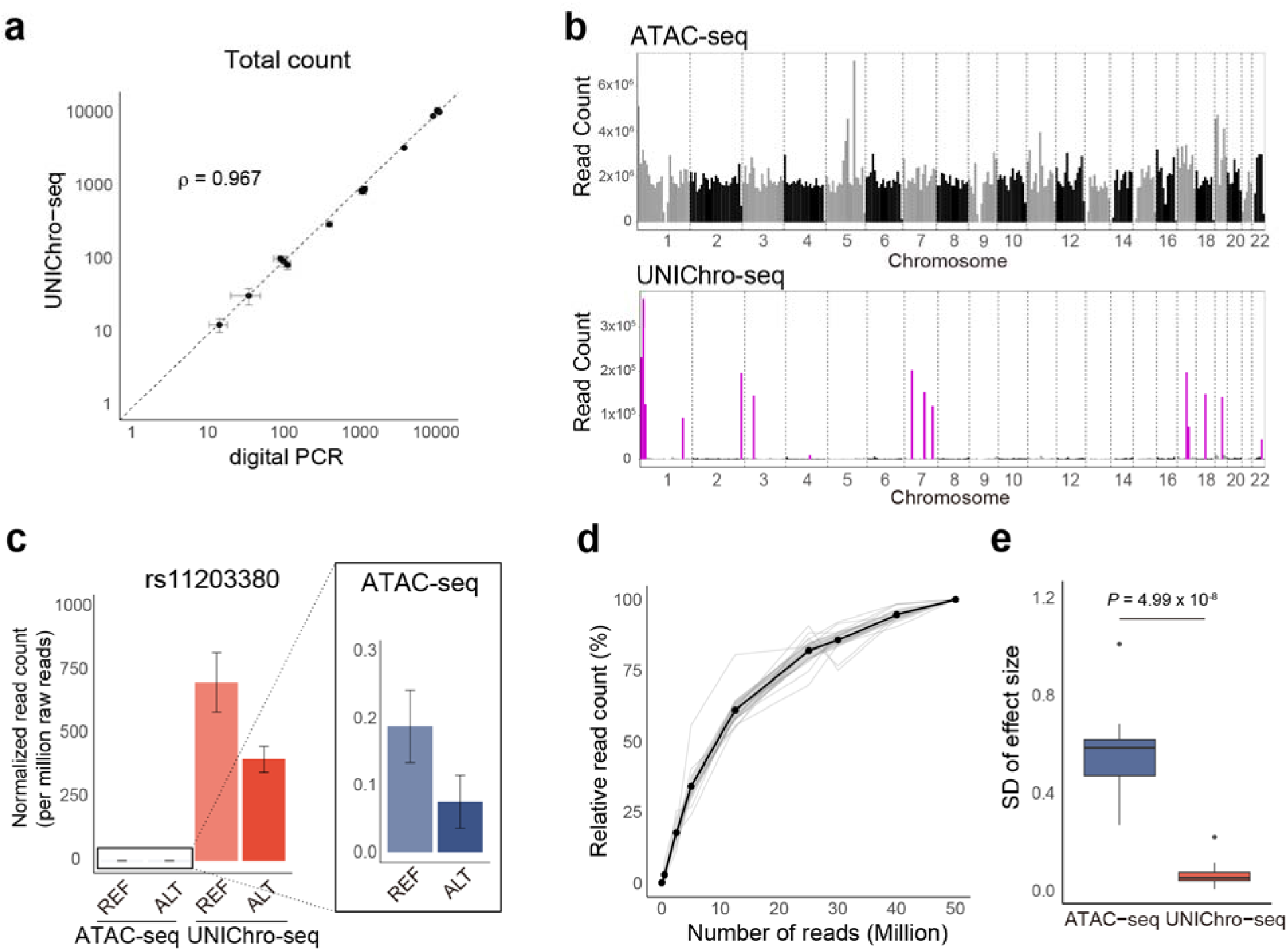
Benchmarking of UNIChro-seq and validation of multi-target UNIChro-seq. **a**, Comparison of the total number of artificial fragments detected by digital PCR and UNIChro-seq. **b**, Enrichment of the target region in conventional ATAC-seq (top) and UNIChro-seq (bottom). The X-axis represents the chromosome position, and the Y-axis denotes the number of mapped reads. The target region is highlighted in purple. **c**, A representative example of the de-duplicated read count normalized to one million raw reads carrying reference and alternative allele of rs11203380 by conventional ATAC-seq (n = 8 replicates) and UNIChro-seq (n = 3 replicates). The error bars represent 95% confidence based on the SEM. **d**, The proportion of de-duplicated reads from conventional ATAC-seq mapped to target loci at different sequencing depths. To visualize all loci (n = 14) on the same scale, read counts at each sequencing depth were expressed as a percentage relative to those at 50 million reads. Each gray line represents an individual target locus. The black line represents the mean of all target loci, and the shaded area represents the 95% confidence interval based on the standard error of the mean (SEM). **e**, The standard deviation of the effect sizes among replicates in conventional ATAC-seq and UNIChro-seq for each target variant (n=14). *P*-value in a Wilcoxon test is provided.

We next sought to apply UNIChro-seq in a realistic scenario (**Supplementary Figure 1a**). Based on the available QTL database, we selected systemic lupus erythematosus (SLE) risk variant (rs2061831^12,22,23^) as a positive control of chromatin-accessibility QTL (caQTL). The UMI count was correlated with the input number of cells (**Supplementary Figure 3b**), indicating that UMIs effectively remove PCR bias even in realistic applications. Using the digital counting of accessible chromatin molecules enabled by UMIs, we estimated that the fraction of cells with open chromatin was approximately 21% (**Methods**). Furthermore, effect size estimates remained nearly constant across different numbers of input cells (**Supplementary Figure 3c**), confirming both the accuracy and sensitivity of the system, which is applicable to as few as 2,000 input cells. These results demonstrated the conceptual viability of UNIChro-seq by highlighting its capacity to assess allelic information at the target region with high accuracy and sensitivity.

### Efficient allelic effect estimation at multiple loci using UNIChro-seq

In a typical scenario, researchers are often interested in multiple variants. Performing UNIChro-seq separately for each variant would waste significant resources, including valuable human samples. This prompted us to evaluate the feasibility of multi-target UNIChro-seq, where we analyze allelic effects at multiple loci simultaneously. We first conducted conventional ATAC-seq with eight replicates of Jurkat cell lines, tested for allelic effects, and identified 17 target loci exhibiting allelic imbalance (**Methods**). A dedicated pipeline was then prepared to design primers for multiple loci to maintain multiplex PCR efficiency while avoiding primer dimers (**Supplementary Figure 4a**). We subsequently performed multi-target UNIChro-seq by including primers for all loci in a single reaction. Although multiplex PCR generally suffers from low amplification efficiency due to primer interference^24^, UNIChro-seq achieved around 18,500-fold enrichment of target loci compared with conventional ATAC-seq (**Figure 2b, c and Supplementary Figure 4b,c**). This result confirms that UNIChro-seq captures substantial molecular complexity, which is critical for sensitive and accurate quantification. Consistent with previous studies^25^, deeper sequencing could not compensate for the insufficient coverage of the target loci in conventional ATAC-seq due to the high duplication ratio, highlighting the importance of target amplification of UNIChro-seq (**Figure 2d** and **Supplementary Figure 4d**). These results confirmed that the conventional approach was inherently noisier than UNIChro-seq due to shallow read depth, low complexity and the lack of UMIs. Therefore, we viewed the results of the conventional approach as reference data with moderate accuracy, rather than as a gold standard. Next, we sought to test the consistency in allelic direction. Among 17 target loci, we excluded three loci during the QC process of UNIChro-seq data (**Methods**). Among the remaining 14 loci, the direction of allelic effect was consistent at 13 loci between both experimental systems (sign test *P*-value = 0.018; **Supplementary Figure 4e**). As expected, the variance in effect sizes across replicates was significantly lower in UNIChro-seq than in the conventional ATAC-seq, confirming the superior accuracy of UNIChro-seq (Wilcoxon test *P*-value = 5.0 x 10^-8^; **Figure 2e**). Together, these results confirmed the feasibility of multi-target UNIChro-seq, demonstrating its superiority over conventional ATAC-seq.

### Investigation on the caQTL effect of immune-mediated disease risk variants

Next, we aimed to demonstrate the utility of UNIChro-seq in variant-to-function (V2F) studies and selected two well-studied complex traits: SLE and rheumatoid arthritis (RA). Their risk variants are enriched in the regulatory regions of B cells and CD4^+^ T cells^2,26–28^, respectively, which we used for this experiment. We first evaluated the replicability of 23 previously reported fine-mapped caQTL detected in a lymphoblastic cell line (LCL; B cell-derived cell line)^29^ that colocalize with SLE risk variants^30^ (posterior probability for caQTL > 0.5; **Methods**). Taking advantage of the efficiency of our system, we collected stimulated B cells derived from five donors at 12 different time points (ranging from 0 to 72 hours post-stimulation) and applied UNIChro-seq (**Figure 3a**). To infer the dynamic nature of caQTL effects in addition to the baseline allelic effects, we included linear and quadratic time terms in our model. Of the 23 loci, 21 had at least two heterozygous donors and were included in the analysis. The baseline effect directions were consistent with those reported in LCL in 17 out of 21 loci (the sign test *P* value = 0.007; **Figure 3b**). Among 12 loci that passed QC, the time-dependent effect was significant at the *ZEP90* locus and suggestive at four other loci (*P* = 1.22 x 10^-9^ and *P* < 0.05, respectively; **Figure 3c**, **Supplementary Figure 5, Supplementary Table 2**). We then sought to discover caQTL effects at 14 fine-mapped risk variants for RA within the accessible regions of CD4^+^ T cells (posterior probability for risk variant > 0.2; **Methods**). We employed the same experimental and analytical protocols, utilizing stimulated CD4^+^ T cells (**Figure 3a**). Among the eight loci with at least two heterozygous donors that passed QC, the baseline effect was significant at seven loci (*P* < 0.05/8; **Figure 3c**, **Supplementary Figure 6, Supplementary Table 3**). This pervasive caQTL effect of the causal variants supports the idea that chromatin accessibility in CD4^+^ T cells mediates genetic risk for RA. The time-dependent effect was significant at the *CD2* locus, which has been well-investigated in previous studies^31,32^ (*P* = 1.02 x 10^-5^; **Figure 3c**). Additionally, we observed a suggestive time-dependent effect for rs58107865 at the *LEF1* locus (*P* = 0.04; **Figure 3c**), and we aimed to validate its dynamic caQTL effect in the following paragraph using a genome-editing experiment. Overall, UNIChro-seq, when specifically targeting candidate causal variants at GWAS risk loci, allows us to assess both steady-state and dynamic allelic effects using a small number of donors.

**Figure 3.**
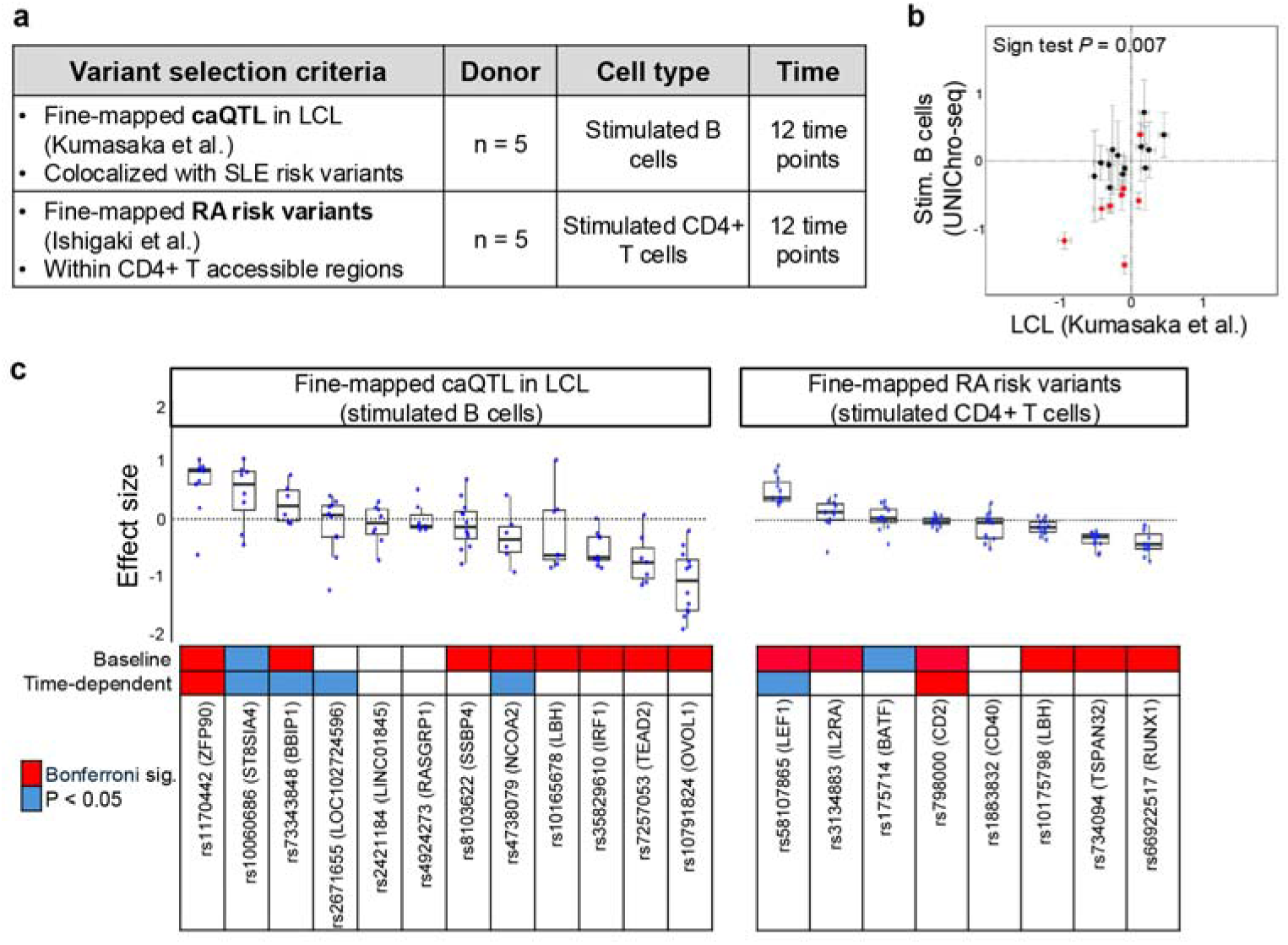
UNIChro-seq successfully validated and identified dynamic caQTL effects of autoimmunity risk variants with primary cells. **a**, Variant selection criteria and sample information of this analysis**. b**, Comparison of the caQTL effect size of 21 SLE risk variants detected in the previous report and our analysis. SLE risk variants with Bonferroni significance in UNIChro-seq analysis are highlighted in red. The sign test P-value is indicated. Error bars represent the 95% confidence interval. **c**, The effect size of the alternative allele at 12 time points is provided for 12 SLE variants and 8 RA variants that passed the quality check filter. Variants with Bonferroni and nominal significance of baseline effect (top row) and time dependency (second row) are highlighted in red and blue, respectively (**Methods)**.

### UNIChro-seq combined with artificial variants introduced through genome editing

Fine mapping of risk variants associated with complex diseases, followed by the experimental validation of their molecular effects using artificially introduced risk variants through precise genome editing, is gaining attention in genetics^19,20,33^. Unlike natural variants, artificial variants are independent of donor-specific haplotype structures, allowing us to infer causal molecular mechanisms at single base pair resolution without being influenced by LD bias (**Figure 1a**). Clearly, UNIChro-seq presents a reasonable option for quantifying molecular effects in this context. However, it is important to note that genetic studies utilizing genome editing are still progressing and may be subject to potential biases in allelic effect evaluation. One major confounder could be the chromatin status. Accessible chromatin regions recruit more CRISPR proteins than closed regions, resulting in higher editing efficiency^34–36^. Conversely, CRISPR proteins and DNA repair enzymes may partially remain at the edited site, keeping the chromatin open^37–40^. These interactions could lead to falsely positive caQTL effects of the edited alleles, which may be dependent on the locus-specific half-life of chromatin accessibility as well as the time elapsed after editing. We refer to this potential bias as “edited-allele bias” (**Figure 4a**). To avoid confusion, we refer to the caQTL effect size that may include the edited-allele bias as “observed effect size” in genome-editing experiments, clearly distinguishing it from the true effect size, which is estimated by excluding the bias.

**Figure 4.**
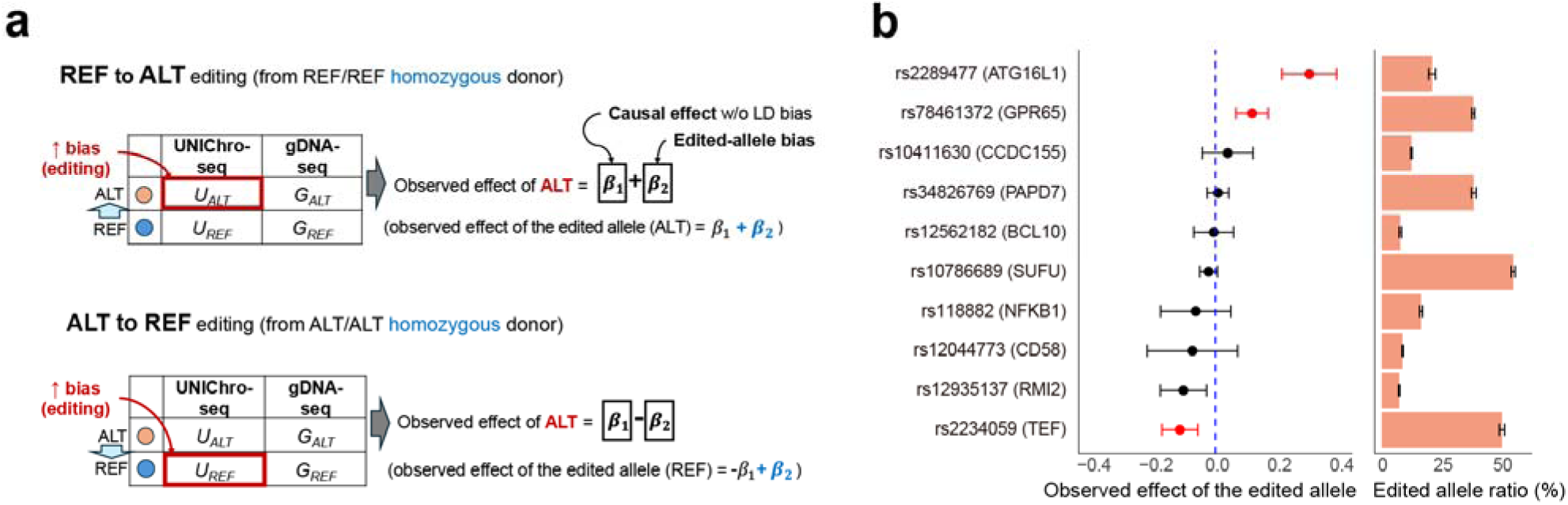
Uni-directional editing and potential edited-allele bias. **a**, Impact of edited-allele bias on effect size estimation. When UNIChro-seq UMI counts exhibit an upward bias for the edited-allele, this bias affects the estimated effect size of the ALT allele in the opposite direction. The observed effect size can be conceptualized as the sum or the difference of the true causal caQTL effect (β₁) and the editing-induced bias (β₂). **b**, Effect size estimates of the edited allele (left panel) and the editing efficiency (right panel). Variants with significant effect sizes are highlighted in red (*P* value < 0.05/10 total tests). The error bar represents the 95% confidence interval from a fitted linear model. For editing efficiency following our analytical procedures using custom in-silico probes, the mean of 11 biological replicates is shown for each variant (n=10).

To assess the feasibility and showcase potential limitations of the approach combining genome editing and UNIChro-seq, we conducted two experiments with uni-directional (an ordinary experimental design) and bi-directional genome editing (our unique experimental design), as described in the following paragraphs. We utilized the latest CRISPR genome editing technology, prime editing 5 (PE5)^41,42^, which allows us to accurately edit variants without introducing double strand breaks. Unlike natural variants in heterozygous donors, artificially edited variants do not exhibit a 1:1 allelic ratio in the genomic DNA. Therefore, we included an offset term for the edited allele frequency obtained from genomic DNA-seq in our model (**Methods**). In both sequencing platforms, we excluded reads with unintended edits before calculating allele frequency using custom-designed in-silico probes (**Supplementary Figure 7a**).

We first conducted uni-directional genome editing at 10 homozygous loci using Jurkat cell lines. To investigate causal allelic effects efficiently, we selected variants with functional evidence from massively parallel reporter assays (MPRA)^43^ and with individual editing efficiencies exceeding 10% using PE5. We edited all loci simultaneously and quantified the edited allele’s caQTL effect two weeks after transfection. Despite the challenges of simultaneous editing, the average efficiency was as high as 25% (**Figure 4b**). This is critical since editing efficiency defines the power to detect QTL effects. Among the 10 loci, the edited alleles of rs2289477 (*ATG16L1*) and rs78461372 (*GPR65*) exhibited a significantly positive observed effect, while rs2234059 (*TEF*) showed a significantly negative observed effect (*P* < 0.05/10; **Figure 4b** and **Supplementary Table 4**). These results suggested that the edited-allele bias does not dominate the findings, and the negative effect at rs2234059 cannot be solely attributed to this bias. However, it is important to note that the uni-directional editing system, being a standard platform, cannot exclude the possibility of bias at each locus, as this bias may be locus-specific. Even for rs2234059, the observed effect could be attenuated by the bias.

To overcome this limitation and distinguish true caQTL effect (β₁) from edited-allele bias (β₂), bidirectional genome editing—introducing both REF-to-ALT and ALT-to-REF edits—serves as an effective solution. In this context, we can reasonably assume that the bias is identical for both editing directions for a given variant (**Figure 4a**), as multiple confounding factors—such as chromosome structure, gDNA break points, and DNA repair mechanisms—are shared. While recruiting multiple donors with both homozygous genotypes and performing bidirectional editing represents the ideal approach, using donors with heterozygous genotypes offers a more practical and feasible alternative due to the allele frequency restriction (**Supplementary Figure 8a**). Additionally, when using a cell line with a single genotype, preparing both homozygous genotypes is not practical. Therefore, we designed a unique experimental platform for bi-directional genome editing from “heterozygous” genotype and employed a logistic regression model to jointly consider the true caQTL effect and the bias (**Figure 5a**). Importantly, this strategy results in a mixture of endogenous and edited alleles, each possessing mutually complementary properties: the endogenous alleles are free from edited-allele bias, while the edited alleles are unaffected by LD bias. To ensure statistical tractability, we approximated the marginal effect size of the endogenous allele with the true caQTL effect (see **Supplemental Note** for potential limitations of this approximation).

**Figure 5.**
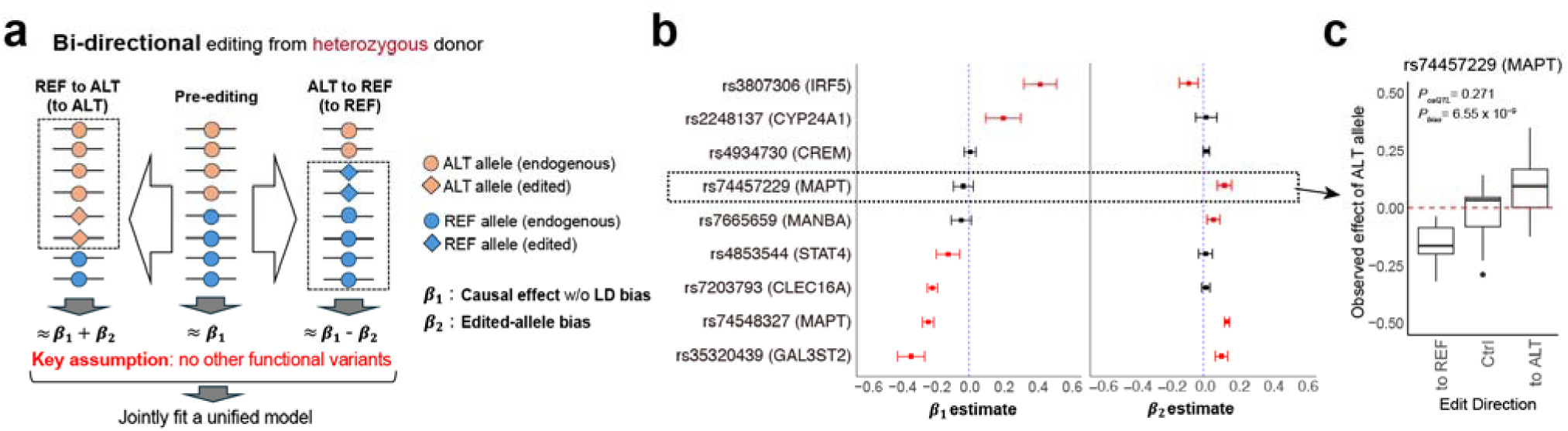
Bi-directional editing estimates the true caQTL effect and the edited-allele bias. **a**, Our strategy of the bi-directional editing using heterozygous samples. Under the key assumption that no other functional variants are present on the same haplotype, the observed effects of the three conditions (pre-editing, ALT-to-REF editing, and REF-to-ALT editing) can be represented as combinations of β₁ (the true causal effect) and β₂ (the edited-allele bias). See the **Supplementary Note** for a detailed discussion. **b**, Effect sizes of the ALT allele (left) and the edited-allele bias (right) at nine loci. We used nine biological replicates for each locus. Variants with significant estimates are highlighted in red (*P* value < 0.05/9 total tests). The error bar indicates the 95% confidence interval from a fitted linear model. **c**, rs74457229 showed a significant edited-allele bias. The Y-axis indicates its observed effect size illustrated in panel **a**.

We selected nine functional variants with heterozygous genotypes in Jurkat cells based on the criteria described above and conducted genome editing in both directions for each variant. The mean editing efficiency was 15.9% for REF to ALT and 15.8% for ALT to REF editing (**Supplementary Figure 7b, c**). Our system successfully differentiated between the true caQTL effect and the bias. Among the nine loci, six demonstrated significant caQTL effects (*P* < 0.05/9; **Figure 5b** and **Supplementary Table 5**). As expected, edited-allele bias was generally positive, with eight loci showing positive effect estimates (binomial test *P* value = 0.039), and five loci exhibited significant edited-allele bias (*P* < 0.05/9; **Figure 5b** and **Supplementary Figure 8b**). Notably, the edited-allele bias at rs74457229 (*MAPT*) was striking, revealing opposing observed effects depending on the direction of editing (**Figure 5c**). Of note, the *IRF5* locus showed a significantly negative edited-allele bias, suggesting that other mechanisms may contribute to the bias. To the best of our knowledge, this represents the first evidence suggesting the existence of edited-allele bias. In conclusion, we demonstrated the importance of bi-directional genome editing, which can be efficiently achieved with prime editing, for evaluating locus-specific QTL effects of edited alleles while distinguishing them from edited-allele bias.

### Functional fine mapping of the RA GWAS variant using UNIChro-seq with artificial variants

Motivated by the robustness of the experimental causal inference enabled by UNIChro-seq combined with genome editing, we next investigated rs58107865, a fine-mapped RA risk variant with the C-allele identified as the protective allele^2^. This is the variant with suggestive evidence of dynamic caQTL (**Figure 3c** and **Supplementary Figure 6**). We edited the rs58107865 genotype in primary CD4^+^ T cells, followed by caQTL effect inference through UNIChro-seq.

We first performed genome editing in homozygous donors. G-to-C editing in five donors homozygous for the G allele (rs58107865 G/G) revealed that the C allele (edited allele) exhibited a positive observed effect three days post-editing (**Figure 6a** and **Supplementary Table 6**). In clear contrast, C-to-G editing in three donors homozygous for the C allele (rs58107865 C/C) showed no observed effect for the C allele at the same time point (**Figure 6a**). These results suggested that the observed signal reflects both a true positive caQTL effect of the C allele and a positive edited-allele bias, with the same magnitude.

**Figure 6.**
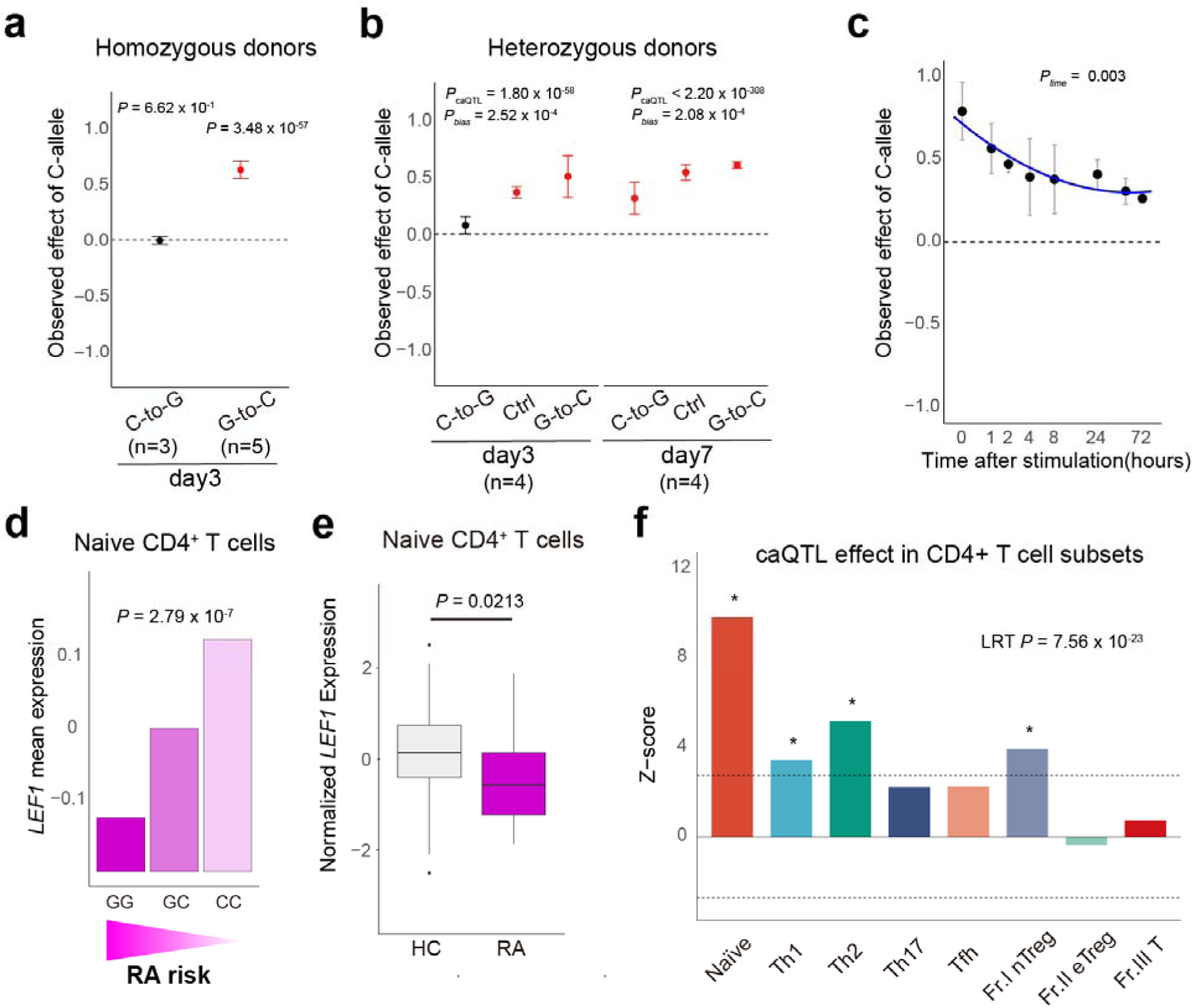
Experimental fine-mapping and functional characterization of rs58107865. **a** The caQTL effect size estimates of the rs58107865-C allele detected by genome editing using homozygous donors for the C-to-G edit (n=3) and the G-to-C (n=5 donors). Significant effects are highlighted in red. b, The caQTL effect size estimates of the rs58107865-C allele detected by bi-directional editing using heterozygous donors (n=4). Significant effects are highlighted in red. *P*-value for caQTL (*P_caQTL_*) and edited-allele bias (*P_bias_*) estimated by a joint model is provided. **c**, The dynamic caQTL effect of the edited C-allele of rs58107865 (n=3 donors). Error bars indicate 95% confidence interval from a fitted linear model. *P*-value for time dependency (*P_time_*) is shown. **d**, The eQTL effect of rs58107865 in naïve CD4^+^ T cells from the ImmuNexUT database (**URL**). **e**, *LEF1* mRNA expression levels in naïve CD4^+^ T cells from healthy individuals and RA patients in ImmunNexUT data. Wilcoxon test *P*-value is provided. **f**, Z-score of the caQTL effect of rs58107865 in CD4^+^ T cell subsets detected by UNIChro-seq (n=6 donors). P-value of likelihood test is provided. *, *P* < 0.05/8. HC, healthy control; RA, rheumatoid arthritis; Naïve, Naïve CD4 T cells; Th1, T helper 1 cells; Th2, T helper 2 cells; Th17, T helper 17 cells; Tfh, T follicular helper cells; Fr.I nTreg, fraction I naïve regulatory T cells; Fr.II nTreg, fraction II effector regulatory T cells; Fr.III T, fraction III non-regulatory T cells.

To confirm the replicability of this observation and further examine the time dependency of the bias, we next performed bidirectional editing in four heterozygous donors (rs58107865 G/C) and monitored the effects through day 7. When the G-allele was edited to C-allele (G-to-C editing), we again detected a positive observed effect of the C-allele on both days 3 and 7. In contrast, after editing the C-allele to G-allele (C-to-G editing), we replicated the attenuated effect of the C-allele, which was more pronounced on day 3 than on day 7 (**Figure 6b** and **Supplementary Table 7**). Consistently, the magnitude of the edited-allele bias on day 7 was 27% less than that on day 3. Our novel platform demonstrated the causally positive caQTL effect of the rs58107865-C allele and highlighted the edited-allele bias, especially shortly after CRISPR perturbation, underscoring the importance of bi-directional editing also in primary cells.

We next sought to confirm the dynamic caQTL effect of rs58107865 using genome-edited CD4^+^ T cells in an eight time-point experiment following stimulation. The analysis successfully revealed a significantly dynamic caQTL effect that continuously attenuated post-activation (**Figure 6c** and **Supplementary Table 8**). Together, our novel experimental platform demonstrated the intriguing molecular effects of rs58107865; it represents a fine-mapped risk variant of RA and a causally dynamic caQTL, with the risk allele leading to decreased chromatin accessibility around the *LEF1* locus. This genetically unique behavior, along with a known immunologically pivotal role of *LEF1* in T cell biology^44–48^, prompted us to further investigate the effect of rs58107865 and *LEF1* in the following paragraphs to understand the RA etiology.

### The molecular effect of rs58107865 in naïve CD4+ T cells affects RA pathology

In the ImmuNexUT database, the largest-scale eQTL study utilizing finely sorted immune cells from patients with immune-mediated diseases, the rs58107865 C-allele (the protective allele for RA) exhibits a significantly positive eQTL effect on *LEF1* expression, specifically in naïve T cells among the eight T cell subsets (**Figure 6d; Method**). The database also revealed that *LEF1* expression levels in naïve T cells are reduced in RA patients compared to healthy individuals (Wilcoxon test *P*-value = 0.02, **Figure 6e**), indicating directional consistency with the genetic risk inferred from the eQTL results.

This observation led us to investigate the caQTL effect in a similar cellular context. We extracted CD4^+^ T cells from five donors with the heterozygous rs58107865 genotype and sorted the same eight CD4^+^ T cell subsets as in the ImmuNexUT database using fluorescence-activated cell sorting (FACS, **Supplementary Figure 9**). Consistent with the eQTL results, UNIChro-seq revealed a significantly positive caQTL effect of the rs58107865-C allele and highlighted its substantial cell type specificity (likelihood ratio test *P* value = 7.56 x 10^-23^). Notably, naïve CD4^+^ T cells exhibited a more significant effect than the other subsets (**Figure 6f** and **Supplementary Table 9**). In conclusion, multiple lines of evidence suggest that the decreased expression of *LEF1*, resulting from reduced chromatin accessibility—particularly in naïve CD4^+^ T cells—is linked to RA pathology.

### Pathogenic genes, pathways, and cell types suggested by *LEF1* silencing

To understand the *LEF1*-mediated risk at a fine scale, we conducted a *LEF1* silencing that mimics the RA genetic risk event, using CD4^+^ T cell samples from three healthy donors. Specifically, we performed *LEF1* knockdown in whole T cells and quantified RNA along with 19 cell surface proteins at the single-cell level using cellular indexing of transcriptomes and epitopes by sequencing (CITE-seq)^49^. Additionally, we purified CD4^+^ T cells to conduct bulk ATAC-seq and RNA-seq. We utilized small interfering RNA (siRNA) to knockdown *LEF1* (*LEF1*-KD) or to target a non-specific control (Ctrl) 24, 48 and 72 hours after transfection.

In the CITE-seq analysis, 37,027 cells remained after standard quality control processes. We applied canonical correlation analysis (CCA) to integrate single-cell RNA and protein information, identifying 13 clusters (**Figure 7a and Supplementary Figure 10a,b**). Among these, three clusters were classified within the naïve CD4^+^ T cell (T_N_) lineage: CD4^+^ T_N_-0 (IL7R^+^), CD4^+^ T_N_-1 (HLA-DR^+^CD38^+^), and CD4^+^ T_N_-2 (TCF7^+^CD45RA^+^). Transcriptomic profiles indicated that CD4^+^ T_N_-0 is in a resting state, whereas CD4^+^ T_N_-1 exhibits an activated phenotype among the T_N_ cells (**Supplementary Figure 10b and Supplementary Table 10**). We first assessed quantitative cellular changes. We tested single-cell level abundance by comparing the frequencies of the 13 clusters between *LEF1*-KD and Ctrl using a generalized linear mixed model. Strikingly, *LEF1*-KD resulted in a 2.3-fold increase in the abundance of CD4^+^ T_N_-1, while it caused a 1.6-fold decrease in the abundance of CD4^+^ T_N_-0 at 48 hours (*P* values = 9.0 x 10^-90^ and 2.1 x 10^-70^, respectively; **Figure 7b**). These cellular effects continued after 72 hours (**Supplementary Figure 10c**). We next examined qualitative cellular changes by assessing differentially expressed genes (DEGs) resulting from *LEF1* knockdown in each cell cluster. Notably, *LEF1* ranked near the top of the DEG list in most clusters, confirming successful silencing. DEGs were substantially shared across cell clusters, particularly within the same lineage (**Figure 7c** and **Supplementary Figure 11a**). Among 3,437 DEGs identified in at least one cell type, 895 DEGs were detected in at least seven of the eight CD4^+^ T cell clusters, including *IRF4*, known to be involved in RA pathogenesis^2,50^ (**Supplementary Table 11, 12**). Given that the T_N_-1 cluster is increased by *LEF1* silencing, we hypothesized that this cluster represents a candidate causal cell type in RA and investigated the characteristics of 1,570 DEGs exclusively found in the T_N_-1 cluster. These DEGs included GWAS candidate genes and cytokine- and chemokine-related genes (**Figure 7d**) and were enriched for T cell activation/differentiation pathways (**Supplementary Figure 11b**).

**Figure 7.**
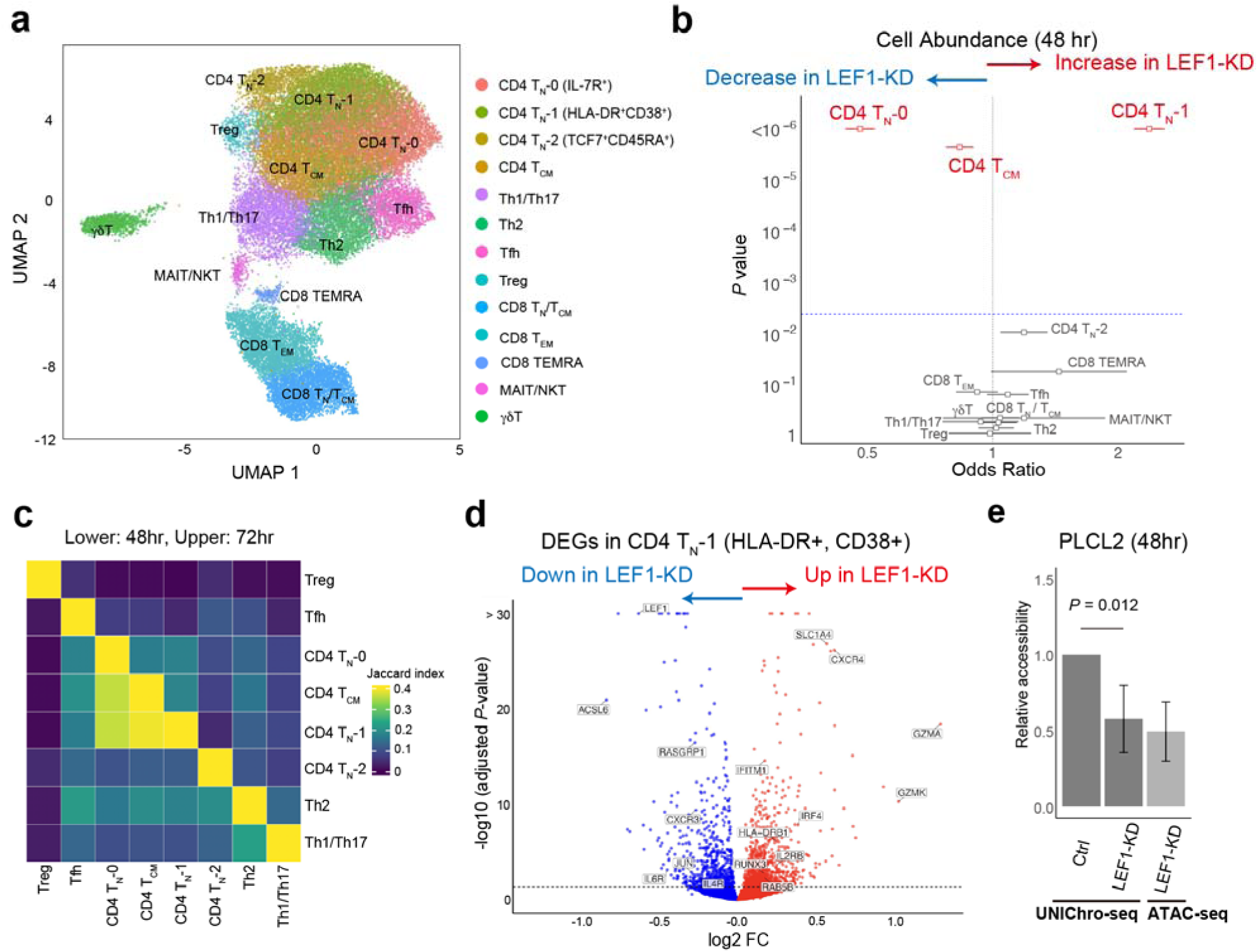
Pathogenic role of *LEF1* in the development of rheumatoid arthritis. **a**, A UMAP embedding of CITE-seq data for *LEF1*-knockdown and control T cells. Colors represent cell clusters (n=13; including eight CD4^+^ T cell lineages). **b**, The shift in the cell type abundance by *LEF1* knockdown estimated by MASC (ref). Cell clusters with a significant shift are highlighted in red. The dashed line indicates the significance threshold (*P* = 0.05/13). **c**, We provided the Jaccard index between the differentially expressed genes (DEGs) of eight clusters in CD4^+^ T cell lineages (FDR < 0.05). **d**, DEGs upon *LEF1* knockdown detected in the CD4 T_N_-1 cluster (HLA-DR+, CD38+). Blue dots indicate downregulated genes and red dots indicate upregulated genes. **e**, Relative accessibility of the *PLCL2* locus compared with the reference locus estimated by UNIChro-seq. *P*-value in Wilcoxon test is indicated.

Bulk ATAC-seq comparing *LEF1*-KD and Ctrl CD4 T cells revealed 56 differentially accessible regions (DARs) associated with *LEF1* silencing (FDR < 0.05; **Supplementary Figure 11c and Supplementary Table 13**). As a sanity check, we evaluated transcription factor binding capacity within the DARs and confirmed that they are enriched for the binding motifs of LEF1 and the TCF family (**Supplementary Figure 11d**), known partners of LEF1^45–47,51^. Intriguingly, all significant DARs exhibited a negative effect, suggesting that LEF1 plays a role in maintaining chromatin accessibility. We focused on the four intersect genes, whose relevance was supported by bulk RNA-seq and ATAC-seq (**Supplementary Table 13**, **14**). Among them, we aimed to replicate the DAR at the *PLCL2* (an RA GWAS candidate gene^2,50^). We digitally quantified chromatin accessibility at these regions and a reference region using UNIChro-seq with the same samples as ATAC-seq (**Method**). We then quantified the relative accessibility of the target regions and successfully replicated the *LEF1*-silencing effect observed in the bulk ATAC-seq with a similar effect size (**Figure 7e, Supplementary Table 15**). In conclusion, we demonstrated that *LEF1* silencing increased the population of activated naïve CD4^+^ T cells and altered chromatin accessibility and gene expression of multiple immune-relevant genes. These molecular events may play a role in the pathogenesis of RA. Additionally, UNIChro-seq enables us to relatively quantify chromatin accessibility in target regions, similar to qPCR.

## Discussion

In this study, we developed a novel method, UNIChro-seq, that digitally counts accessible chromatins in target regions, and we extensively demonstrated its advantages over conventional ATAC-seq. The UMI-based counting approach enables accurate digital counting, while the target amplification method successfully enriches target regions, thereby improving sensitivity and reducing the sequencing cost substantially (e.g., approximately 18,500-fold reduction in multi-target UNIChro-seq). Moreover, its efficiency allows an expanded search space of cellular states for QTL detection. When combined with genome editing, UNIChro-seq becomes a powerful tool for inferring causal molecular effects and their dynamic behavior.

Recently, statistical fine-mapping of disease risk variants has emerged as a major trend in genetic research, driven by multi-ancestry study designs and sophisticated statistical models. Therefore, the field requires a toolbox for experimental fine-mapping, and UNIChro-seq is an ideal tool for this purpose. We showcased the power of UNIChro-seq using edited variants at 20 loci in total, demonstrating the causal QTL effect at 10 loci. Among these, the most successful example is rs58107865, a fine-mapped GWAS variant associated with RA at the *LEF1* locus.

The edited allele bias, defined as the spurious molecular effect of the edited alleles, is a concept we first demonstrated in this study. We identify several characteristics of this bias. First, the bias can be sufficiently large to obscure the true causal QTL effect (**Figure 6a**). Second, this bias is locus-specific; thus, not observing an effect at one locus does not guarantee that the system is free from bias at other loci. Third, this bias is likely to be prominent shortly after the editing occurs. Since chromatin accessibility correlates with multiple molecular phenotypes, including gene expression, the bias should not be limited to caQTL. Furthermore, it is likely not specific to the prime editing used in this study and may confound other CRISPR-related technologies. Employing bi-directional editing offers a robust solution for distinguishing the bias from the true allelic effect. Therefore, we urge the community to pay close attention to edited-allele bias and to rule it out when suspected by using bi-directional editing. To achieve this aim, the best practice is to recruit donors with both homozygous genotypes. However, when this is not feasible, using a heterozygous donor may offer an alternative solution (see **Supplementary Note** for further discussion).

There are several limitations to our study. First, the success of target enrichment depends on the quality of nested primers in the target region. Some regulatory regions have excessively high GC content, which can complicate primer design. Second, we observed reduced efficiency in multi-target UNIChro-seq. Variations in accessibility across targets may create a significant imbalance in the number of reads mapped to target regions, making it challenging to obtain sufficient UMIs in less accessible areas. A potential solution is to group the target regions into several categories based on their level of chromatin accessibility. Lastly, although infrequent, we noted several unintended edits in prime editing. We carefully designed the analytical pipeline to remove sequence reads originating from these unintended edits, making our analysis largely robust against such occurrences. However, we cannot completely rule out these effects since we cannot evaluate mutations outside the sequencing range.

In summary, our novel experimental strategy that utilizes UNIChro-seq can be applied to various experimental scenarios, particularly when investigating edited alleles created by CRISPR technologies. Our system contributes to advancing V2F research and deepens our understanding of disease etiology.

## Online Methods

### Sample collection and processing

LCL cell lines (ND11920) and Jurkat E6-1 cell lines (88042803) were obtained from the Coriell Institute and ECACC, respectively. Whole blood from healthy individuals was obtained with informed consent and approval by the institutional review boards. Peripheral blood mononuclear cells (PBMCs) were isolated with Lymphoprep density gradient medium (STEMCELL, ST-07801) and stored at -150°C in CryoStor CS10 (Sigma, C2874-100ML). Human primary CD3^+^ T cells, CD4^+^ T cells, and B cells were isolated from freshly thawed frozen PBMCs using MACS (Myltenyi, 130-096-535, 130-096-533 and 130-101-638, respectively). We sorted PBMCs into eight CD4^+^ T cell subsets with purity > 95% using a FACS Aria III (BD Biosciences) with the flowcytometry staining panel reported in the ImmuNexUT^12^ (**Supplementary Figure 9 and Supplementary Table 16**). We targeted 20,000 cells, with at least 2,000 per subset. Sorted cells were immediately centrifuged and stored in CryoStor CS10 (Sigma-Aldrich, C2874-100ML) for UNIChro-seq.

Genomic DNA was automatically extracted with the EZ1&2 DNA Tissue Kit (QIAGEN, 953034) using EZ2 Connect (QIAGEN), and RNA was isolated using the MagMax-96 Total RNA Isolation Kit (Thermo Fisher Scientific, AM1830). All the cell culture conditions are explained in the **Supplementary Note**.

### Prime editing

All the sequences of pegRNAs, epegRNAs, gRNAs and sgRNAs used in this work are listed in **Supplementary Tables 17**, and our strategy for designing them are explained in the **Supplementary Note**. mRNAs encoding PEmax and hMLH1dn were prepared by in-vitro transcription (IVT) using the protocol described previously^41,53^. Briefly, the mRNA transcription template was generated by PCR with pT7-PEmax for IVT and pT7-hMLH1dn for IVT (Addgene #178113 and #178114). mRNA was generated by IVT using a HiScribe T7 High-Yield RNA Kit (NEB, E2040S). All mRNA was produced with UTP fully replaced with N1-methylpseudouridine-5’-triphosphate (TriLink, N-1081) and co-transcriptional capping by CleanCap Reagent AG (TriLink, N-7113). The transcribed mRNA was purified through lithium chloride precipitation and diluted with TE Buffer (Nippon Gene 314-90021).

For prime editing of Jurkat cell lines, we pooled the pegRNA and gRNA expression vectors for all loci. In bi-directional editing, vectors for REF to ALT editing and ALT to REF editing were separately mixed. Jurkat cells were electroporated using Neon transfection system 10 μl kit (Thermo Fisher) with 3 x 10^5^ cells resuspended with Buffer R, 750 ng pCMV-PEmax-PE2-GFP (Addgene Plasmid, #180020), 500 ng pegRNA expression plasmids (without in control samples), 168 ng sgRNA expression plasmids and 750 ng MLH1dn plasmid in total of 10 μl per sample with the following conditions; 1350mV, 30ms, three pulses. Cells were plated in pre-warmed media as described above. One day after transfection, GFP-positive cells were sorted out with BD FACS Melody (BD Biosciences) and cultured for 14 days before genomic DNA extraction.

For prime editing in CD4^+^ T cells, prior to electroporation, cells were activated for 2 days with Dynabeads Human T-Activator CD3/CD28 (Thermo, DB11132) at a 1:1 bead: cell ratio and cultured in T cell media (CTS T cell Expansion SFM (Thermo A1048501) supplemented with 2.5% CTS Immune Cell SR (ThermoFisher, Waltham, MA), 200 μM L-Glutamine (Wako, 073-05391), 100U/mL Penicillin/Streptomycin (Thermo Fisher, 15140122)) in the presence of 50 IU/mL IL-2 (Peprotech, 200-02), 5 ng/mL IL-7 and IL-15 (Peprotech, 200-07, 200-15) at 37 °C and 5% CO2. CD3/CD28 beads were removed from cells 5 h before electroporation. For electroporation, 3.0 x 10^5^ cells per sample were pelleted by centrifugation for 5 min at 300 g and resuspended in Buffer T, followed by addition of 2 μg PEmax mRNA, 90 pmol synthetic pegRNA, 60 pmol and synthetic sgRNA, and 2 μg MLH1dn mRNA. Electroporation was performed using 10 μL Neon tips with the following parameters: 1,400 V, 10 ms, three pulses. Cells were plated in 1 mL fresh T cell media supplemented with 2 ng/mL IL-7 and IL-15 (Peprotech) in a 24-well plate. T cells were maintained at around 1 × 10^6^/mL and at 3 and 7 days after transfection 3-5 x10^5^ cells were pelleted by centrifugation for 5 min at 300 g for genomic DNA isolation and 20,000 cells were stored in Cryostor CS10 (Sigma, C2874-100ML) for UNIChro-seq experiments.

To evaluate editing efficiency, edited sites were amplified from genomic DNA templates by nested PCR with the same reverse primers as UNIChro-seq. Libraries were sequenced on an Illumina MiSeq. Universal primer and gene-specific primer sequences for gDNA-seq library preparation are available in **Supplementary Table 18 and 19**.

### Analytical strategy of DNA-seq

The common analytical processes of DNA-seq are outlined in this paragraph. Adapter sequences were trimmed using Cutadapt^54^ (v2.6). We mapped the trimmed reads to the GRCh38 reference genome using Bowtie2^55^ (v2.2.6). After mapping, we filtered the reads using samtools^56^ (v1.9) with flags “-f 2 -q 30” to retain only properly paired reads with high mapping quality. For target variant analysis, we extracted reads within ±300bp of each variant position. For allele-specific analysis, we separated reads containing reference (REF) and alternative (ALT) alleles using custom in-silico probes designed for each variant. We then counted the number of reads supporting each allele to quantify allelic ratios at target sites.

### Experimental strategy of UNIChro-seq

All the sequences of UNIChro-seq nested-PCR primers for each target locus are listed in **Supplementary Table 19**, and our strategy for designing them are explained in the **Supplementary Note**.

We designed the custom transposome adapters as previously reported with some modifications^57^. Each of the mosaic end (ME)-A adapters contains 19-bp ME sequence in the 3’ end for the binding of Tn5, 8-bp barcode sequence to pool samples, a 17-bp UMI sequence, and standard or modified TruSeq sequencing adapter in the 5’ end (**Supplementary Figure 1b**). ME-A and ME-B adapters were first annealed to MErev, respectively. Annealed ME-A and B adapters were mixed in a 1:1 ratio and then assembled with Tagmentase (Diagenode) for 30 min at 23°C according to the manufacturer’s protocol. After adding half a volume of glycerol, the transposon complex was stored at -20 °C. The oligonucleotides and primers used for UNIChro-seq were synthesized in Eurofin Genomics.

After washing and pelleting the cells, we carefully removed the supernatant and resuspended with custom transposome reaction mix (1.25 μl transposome, 12.5 μL 2x tagmentation buffer, 0.25 μl 15 digitonin and 6 μl ddH_2_0). The transposition reaction was performed at 37 °C for 30 min, then the DNA was purified using Zymo DNA Clean & Concentrator-5 (D4014) and eluted in 20 μL resuspension buffer (RSB; 10mM Tris-HCL pH8.0). UNIChro-seq library was constructed with 10 μL of tagmented DNA by linear amplification and semi-nested PCR with 3’-phosphorylated blocking primers with the same sequence as ME to prevent the UMI overwriting by the free adapter or PCR chimera. First, linear amplification was performed with 5’ biotinylated gene-specific primer (outer primer) and the products were pulled down using Dynabeads M280-streptavidin beads. Second, the target region was amplified by the 1^st^ round PCR with the inner primer and the primer designed in the transposome tag sequence, followed by the purification with AMPure beads to select the range from 150 to 1000bp. Finally, we conduct the 2^nd^ round PCR using primers containing adaptor and index sequencing compatible with the Illumina platform. From the 2^nd^ round PCR, products were attached with different barcodes and can be pooled. Final libraries are sequenced on the NovaSeq 6000 platform using a custom R1 sequencing primer. Oligonucleotide and primer sequences are available in **Supplementary Table 20 and 21**.

### Analytical strategy of UNIChro-seq

The common analytical processes of UNIChro-seq are outlined in this paragraph. We de-multiplexed sequence reads using the barcode attached to the Tn5 adaptor, trimmed the adaptors with Cutadapt, and mapped the reads to the reference genome by Bowtie2. In this study, we utilized the GRCh38 genomic coordinate. After mapping, we filtered the reads using samtools with flags “-f 2 -q 30”. For a given variant, we extract the reads overlapping the target variants and split the reads into those with the REF allele and those with the ALT allele using custom-designed in-silico probes. Importantly, the probe is long enough to exclude the reads harboring unintended edits. As a QC process of UMI, we merged the group of UMIs within the Hamming distance of 2 or less. If the same UMI was detected in both the REF and ALT reads, in both target and non-target regions, and in multiple sample indices with same barcode, we excluded the UMI from the group with the lower number of detections to account for the possibility of UMI hopping during library preparation. Furthermore, we set a minimum cutoff as the QC filter from the histogram of UMI detection counts. (**Supplementary Figure 2b**). We treated individual UMI as an independent observation and quantified the number of UMI for the REF and ALT alleles separately to estimate allelic imbalance. Assuming the Tn5 tagging efficiency is 100%, the total UMI count for a given variant indicates the number of accessible chromatin molecules. Therefore, we estimated the fraction of cells with open chromatin by dividing the total UMI count by the theoretical total chromatin molecule.

In the analyses illustrated in **Figure 2**, we aimed to identify variants showing the allelic imbalance in chromatin accessibility and conducted eight biological replicates of conventional ATAC-seq using Jurkat cell line. We mapped the sequence reads to the reference genome by Bowtie2, removed duplicated reads by Picard (v2.26.2) and Samtools, excluded blacklisted regions obtained from ENCODE blacklist for hg38 (see URLs), and called narrow peaks by MACS2^58^ (v2.2.7.1). We called the variant genotypes and the read coverage supporting each allele from ATAC-seq data using Bcftools^59^ (v1.12). As a quality control, we summed up the reads supporting each allele separately and finally included 4,265 biallelic variants where both sums are more than 10 reads. In this analysis, the depth of the target loci was much shallower than UNIChro-seq since the reads were not enriched, and we treated deduplicated reads by Picard (i.e. deduplication based on the mapped position information without UMI information) as an independent event. Therefore, the analysis was inherently noisier than UNIChro-seq and hence we treated these results as reference data with moderate accuracy, rather than the gold standard. To quantify allelic imbalance, we fitted a mixed effect logistic regression model to the ATAC-seq allelic dosages, incorporating binary indicators for each variant as fixed effects and sample-specific factors across all eight replicates as random effects. This approach allowed us to control for general mapping bias. After applying an FDR < 0.1 threshold, we identified 27 variants with significant allelic imbalance. From these, we selected 17 variants for which we could successfully design primers for UNIChro-seq. For each variant, we conducted three biological replicates of UNIChro-seq. In the QC process of UNIChro-seq, we excluded three loci: two loci were not heterozygous in UNIChro-seq (suggesting the genotyping error of bulk ATAC-seq), and 1 locus for poor mappability. We then included 14 loci for the downstream analyses. For these 14 loci, we performed read counting in both UNIChro-seq and conventional ATAC-seq and compared their effect sizes.

In the analyses illustrated in **Figure 3**, we applied UNIChro-seq to infer dynamic shifts in the risk alleles’ QTL effects of RA (using stimulated CD4^+^ T cells) and SLE (using stimulated B cells) across 12 different time points. We selected fine-mapped caQTL variants in tight LD with SLE risk variants using the following approach. We utilized the fine-mapped caQTL variants detected in immortalized B lymphocytes derived from a European population (Kumasaka et al^29^.; posterior probability > 0.5) and the lead variants in GWAS of SLE conducted in EAS populations^30^. Since the full summary statistics were not available for GWAS, we evaluated the colocalization based on the linkage disequilibrium (LD; *r*^2^ > 0.5 both in EUR and EAS). By these steps, we identified 26 caQTL variants and confirmed all are within the accessible region in immortalized B lymphocytes. Among them, we were not able to design primers for UNIChro-seq for three variants, and we included 23 variants for the downstream analysis. We calculated *r*^2^ using the samples in the 1000 Genomes Project (phase 3 version 5). Additionally, we included 12 fine-mapped RA GWAS risk variants (posterior probability > 0.2), that are located within the accessible region of stimulated CD4+ T cells. For each variant, we fitted a mixed effect logistic regression model as follows:

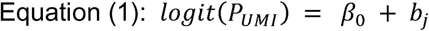

where *P*_UMI_ represents the ALT allele ratio calculated from UMI counts in UNIChro-seq, *β* _0_ represents the fixed caQTL effect, and *b*_j_ is a random intercept term for the *j*-th donor.

For time-course analysis, we filtered out data points with fewer than 6 UMIs in either reference or alternative allele counts. We retained only time points with at least two donors after filtering and excluded variants with fewer than six remaining time points. This ensured high-quality measurements with sufficient donor replication and temporal coverage. We fitted a mixed effect logistic regression model incorporating quadratic time terms as follows:

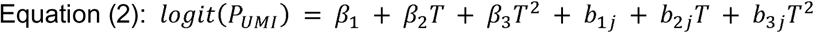

where *P*_UMI_ represents the ALT allele ratio calculated from UMI counts in UNIChro-seq, and *T* and *T*^2^ denote the log-scaled time and its squared value, respectively. *β_1_* represents the fixed caQTL effect, while β*_2_* and β*_3_*capture the linear and quadratic components of the dynamic shift in effect size of the ALT allele on chromatin accessibility over time. *b_1 j_*, *b_2_* _j_ and *b_3_* _j_ are random effect terms for intercept, linear time, and quadratic time components, respectively, accounting for donor-specific variation from the *j*-th donor.

For clarity, we refer to Equation (1) as Model 1 and Equation (2) as Model 2 throughout the manuscript and supplementary materials.

In the analyses illustrated in **Figure 4** and **5**, we referred to the previously reported MPRA results using Jurkat cell lines and extracted 313 functional variants showing evidence of allelic imbalance in gene expression regulation^43^. Among them, we included variants within accessible chromatin regions and chose the variants whose genotype of Jurkat cell line is homozygous for uni-directional editing and those whose genotype is heterozygous for bi-directional editing. We excluded variants with low editing efficiency (< 10%) and finally included 10 homozygous variants and nine heterozygous variants. We conducted 11 biological replicates of UNIChro-seq for uni-directional editing and nine biological replicates for bi-directional editing. We fitted a mixed effect logistic regression model as follows (equation 3 for uni-directional editing and 4 for bi-directional editing):

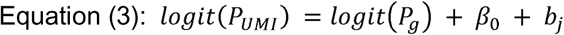

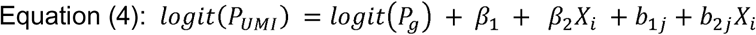

where *P_UMI_* indicates the ALT allele ratio calculated by UMI of UNIChro-seq, *P_g_* is an offset term indicating the ALT allele ratio calculated by reads count of genome DNA-seq. In equation (3), *β_0_* is a fixed effect intercept term indicating the effect size estimate on the chromatin accessibility, and *b_j_* is a random intercept term for the *j*-th replicate sample. In the equation (4), we replaced *β_0_* in the equation (3) by *β_1_* and *β_2_ X_i_*.

*β_1_* is a fixed effect intercept term indicating the true effect size estimate on the chromatin accessibility controlling the edited-allele bias. *β_2_* is a fixed effect coefficient indicating the effect size estimate of the edited-allele bias, where *b_1j_* is a random intercept and *b_2j_* is a random slope for *X_i_* in the *j*-th replicate sample, and *X_i_* is a three-level variable indicating the editing direction: 0 for no editing, 1 for REF to ALT editing, and -1 for ALT to REF editing.

In the analyses illustrated in **Figure 6**, we conducted genome editing of rs58107865 using CD4^+^ T cells from five homozygous donors (uni-directional editing) and from four heterozygous donors (bi-directional editing) and conducted UNIChro-seq at day 3 and 7. We utilized equation (3) for uni-directional editing and (4) for bi-directional editing. Additionally, we performed time course analysis to investigate temporal changes in chromatin accessibility using equation (2). For caQTL analyses in the fine-sorted immune cell, we fitted a mixed effect logistic regression model as follows:

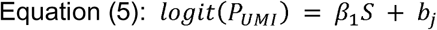

where *P_UMI_* indicates the ALT allele ratio calculated by UMI of UNIChro-seq, and *S* is a binary variable indicating whether the data belongs to the target cell type or not. *β_1_* indicates the gap in the effect size of the ALT allele on the chromatin accessibility between the target cell type and the others. *b_j_* is a random intercept term indicating the *j*-th donor. We performed a likelihood ratio test (LRT) to assess the statistical significance of the cell type-specific effect by comparing this model with a null model without the fixed effect term *β_1_S*.

### Quantitative ATAC-seq (qATAC) assay

We developed a quantitative ATAC-seq (qATAC) assay to precisely measure chromatin accessibility changes at specific genomic loci. We utilized previously identified differentially accessible regions (DARs) from bulk ATAC-seq data in a LEF1 knockdown model. Primary CD4+ T cells were isolated from three healthy donors. For each donor, cells were divided into control (siCT) and LEF1 knockdown (siLEF1) conditions and harvested at 48h post-transfection. We targeted two genomic regions: one that showed significant accessibility changes in response to LEF1 knockdown in bulk ATAC-seq (chr3:16900771-16902017 near PLCL2 gene), and a reference region that exhibits stable accessibility in stimulated T cells (chr5:139560809-139562343 near UBE2D2 gene), commonly used as a reference gene in qPCR.

For each experimental condition, we prepared three technical replicates. To minimize batch effects, consistent barcodes were used across all conditions for each donor. Probes were designed to avoid common SNPs, and the assay was optimized to measure only reference allele counts. These qATAC experiments were performed using the UNIChro-seq method, which enables highly sensitive detection of chromatin accessibility at specific genomic loci. For comparison, we utilized bulk ATAC-seq data from a separate LEF1 knockdown experiment performed in CD4+ T cells.

For data analysis, we calculated the ratio of target region counts to reference region counts for each sample like the ΔCt method in qPCR. Technical replicate means were calculated for each donor and condition. Values were then normalized to corresponding control samples to obtain relative accessibility measurements. Statistical significance was assessed using generalized linear mixed models with Bonferroni correction for multiple testing.

### Artificial fragments for the UNIChro-seq validation

Target region including rs2061831 was amplified by PCR with genomic DNA isolated from LCL (ND11920) which has heterozygous genotype of rs2061831. We confirmed that rs2061831-C and T were 1:1 by Sanger sequence (Eurofin). The PCR products were digested to 259-bp and 60-bp fragments using Scf-I (NEB) according to the manufacturer’s protocol, followed by purification with AMPure beads to select the longer fragment. Oligonucleotides with the same component of transposon adapter with 5’ overhang of TGCA (Eurofin) were annealed and ligated to the digested longer fragment with T4 Ligase. Final products were purified by BluePippin to select at around 320 bp. The number of the artificial fragments and rs2061831 allelic ratio were quantified by QIAGEN QIAcuity digital PCR system using QIAcuity Nanoplate 8.5K (QIAGEN, 250021), QIAcuity Probe PCR kit and TaqMan SNP genotyping assay kit (Thermo Fisher, Assay ID C_1886941_20) according to the manufacturer’s protocol. The sequences of oligonucleotides and primers are listed in **Supplementary Table 22**.

### *LEF1* knock down experiment

Small interfering RNA (siRNA) specific to *LEF1* and control siRNA were purchased from Thermo Fisher. Before transfection of siRNA, CD3^+^ T cells isolated from 3 healthy donors were activated for 2 days with Dynabeads Human T-Activator CD3/CD28 (Thermo, DB11132) at a 1:1 bead: cell ratio and cultured in T cell media supplemented with 30 U/ml IL-2. 3.0 x 10^5^ cells per sample were resuspended in 10 μL NEON T-Buffer, 100 pmol of each siRNA against *LEF1* or 100 pmol control siRNA was added, and cells were electroporated using the following program: 1,400 V, 10 ms, three pulses. The transfected cells were rescued immediately after transfection in pre-warmed T cell media supplemented with 30 U/mL IL-2. The cells were harvested for UNIChro-seq (20,000 cells/sample) and CITE-seq (3 x 10^5^ cells/sample) at 48- and 72-hours post-silencing and, RNA-seq (1 x 10^5^ cells/sample) and ATAC-seq (20,000 cells/sample) at 24-, 48- and 72-hours post-silencing. RNA-seq and ATAC-seq libraries were sequenced on the NovaSeq 6000 Illumina platform, while CITE-seq libraries were sequenced on the NovaSeq X Plus.

For CITE-seq analysis, we pooled the cells from 3 donors for each time point (48h and 72h). We stained up to 300,000 cells per one condition. After Fc receptor blocking with Human TruStain FcX (BioLegend, 422302), cells in each condition were stained with a mixture of a total of 19 TotalSeq-C antibodies including TotalSeq-C anti-human Hashtag antibodies to distinguish between siLEF1 and control condition (BioLegend; **Supplementary Table 23**) for 30 min at 4 °C. We prepared gene expression (GEX) and antibody-derived tag (ADT) libraries using the Chromium Next GEM Single Cell 5’ Kit v2 (10x Genomics) following the manufacturer’s protocol. We processed the sequence data with Cell Ranger 7.0.1^60^ (10X Genomics) and analyzed the outputs using a standard pipeline from the R package Seurat^61^ (v4.4.0). Briefly, we normalized the GEX data with log-transformation (each gene expression count divided by the total expression in each cell multiplied by a scale factor of 10,000 and added one pseudo-count then natural-log transformed) and normalized the antibody-derived tag (ADT) data with centered log-ratio (CLR). We integrated GEX and ADT data through canonical correlation analysis (CCA) using the 2,891 genes (excluded mitochondrial genes, chromosome Y genes, and cell cycle genes from the top 3,000 highly variable genes) and 19 ADTs, following a strategy reported in a previous study^62^. After batch correction of canonical variables with the R Package Harmony^63^ (v1.0.3), we constructed a nearest neighbor graph utilizing the top 11 canonical variables and performed clustering and dimensional reduction through uniform manifold approximation and projection (UMAP). To infer the cell type abundance shift by *LEF1*-KD, we applied MASC^64^. Briefly, we created a binary vector that indicates whether each cell belongs to a target cell type cluster, and fed that vector into a mixed effect logistic regression model where the intervention status (si-RNA or Ctrl) and cell level covariates (the number of UMI and the ratio of reads from mitochondria genes) are the fixed effect explanatory variable and donor and batch IDs are the random effect explanatory variables.

For bulk analysis, RNA-seq libraries were prepared with SMART-seq mRNA (Takara Bio, 634773) and Nextera XT DNA Library Prep Kit (illumina, FC-131-1096). We mapped the sequence reads to the reference genome by STAR^65^ (v2.7.9a) and quantified the reads within the gene region by RNA-SeQC2^66^ (v2.4.2). We utilized the gene model from GENCODE^67^ v26. DEGs were detected by DESeq2^68^ (v1.34.0). We performed ATAC-seq as previously described with a minor modification. Briefly, 200,000 cells were washed 200 μl cold 1xPBS twice and centrifuged at 500g for 5mins. The tagmentation reaction was carried out by adding 1.25 μL TDE1, 12.5 μL 2x TD buffer (illumina, 20034198), 0.25 μL 1% digitonin (Promega G944A) and 6 μL ddH_2_O. The tagment DNA was purified using Zymo DNA Clean & Concentrator-5 (D4014) and amplified using KAPA HiFi HotStart ReadyMix (NIPPON Genetics, KK2602) and Nextera XT index kit v2 (illumina) for 10-12 cycles of PCR. The libraries were purified with AMPure beads to select the region between 180-1000 bp. ATAC-seq data were analyzed following the methods described for Figure 2 in the ’Analytical strategy of UNIChro-seq’ section of Methods.

## Data availability

All raw sequence data from single-cell and bulk RNA-seq and ATAC-seq analyses are available at the Japanese Genotype-phenotype Archive (JGA, https://www.ddbj.nig.ac.jp/jga), which is hosted by the Bioinformation and DDBJ Center, under the accession number JGAS000818.

## URL

ImmuNexUT: https://www.immunexut.org BioRender: https://www.biorender.com/ ENCODE hg38 blacklist: http://mitra.stanford.edu/kundaje/akundaje/release/blacklists/hg38-human/hg38.blacklist.bed.gz

## Code availability

The code and analysis pipeline for UNIChro-seq are available at https://github.com/hhat/UNIChro-seq-Analysis/ and https://doi.org/10.5281/zenodo.15892172.

## ACKNOWLEDGEMENT

This work was supported by funding from the Japan Agency for Medical Research and Development (AMED) (JP22tm0424223, JP22ek0410099, JP23tm0524005, JP25ek0410139, JP22ama121015 and JP223fa627010), JSPS Grant-in-Aid for Scientific Research (B) (22H03114) and Grant-in-Aid for Research Activity Start-up (21K20647), Grant-in-Aid for Early-Career Scientists (25K19621), Keio University Academic Development Funds, Program for the Advancement of Next Generation Research Projects, GSK Japan Research Grant 2021, Daiichi Sankyo Foundation of Life Science, Mochida Memorial Foundation for Medical and Pharmaceutical Research, The Uehara Memorial Foundation, and Takeda COCKPI-T Funding. The super-computing resource was provided by Human Genome Center, Institute of Medical Sciences, The University of Tokyo and RIKEN. We also acknowledge Drs. Yuriy Baglaenko, Soumya Raychaudhuri, and Gregory Newby for helpful advice on genome editing technology. We appreciate the RIKEN-IMS Genome Platform for their help in the sequencing experiment for this study.

## AUTHOR CONTRIBUTIONS

M.K. and K.I. conceived and designed the study. M.K conducted all experiments with support from R.B., T.K., M.S., H.I., T.N.,and B.N.. H.H. conducted all analyses except single-cell analysis with support from M.K., M.N., T.I., H.T. and K.I. K.A. conducted single-cell experiments and analyses with support from M.K., H.H., H.I., T.N. and M.N.. A.S. and K.Y. provided human samples for the investigation of caQTL effect of RA and SLE risk variants. M.K., H.H. and K.I. wrote the manuscript with critical inputs from all co-authors.

