## Supplementary Notes and Figures for "Accurate, sensitive, and efficient chromatin accessibility quantification at target loci using UNIChro-seq"

**Supplementary Note**

**Potential limitations for bi-directional genome editing from heterozygous donors**

Bidirectional editing provides a robust approach for disentangling the true allelic caQTL effect (β₁) from the edited-allele bias (β₂). To achieve this, the optimal strategy is to recruit donors with both homozygous genotypes. In this design, one allele is edited while the other remains endogenous, allowing the complete elimination of linkage disequilibrium (LD) structure bias. As a result, the observed effect for the ALT allele corresponds to β₁ + β₂ when the ALT allele is edited, and β₁ − β₂ when the REF allele is edited (**Figure 4a**). Thus, this approach not only enables the separation of true caQTL effects from editing-induced bias but also facilitates experimental fine-mapping.

However, as shown in **Supplementary Figure 8a**, recruiting donors with both homozygous genotypes can be challenging due to allele frequency constraints. For instance, if the target allele has a frequency of 1%, approximately 10,000 donors would be required to identify a single homozygous individual. To address this limitation, we sought to develop an alternative strategy using “heterozygous” donors. In bidirectional genome editing with heterozygous donors, both endogenous REF and ALT alleles are retained across all three conditions (pre-editing, ALT-to-REF editing, and REF-to-ALT editing). This results in a mixture of endogenous and edited alleles, each possessing mutually complementary properties: the endogenous alleles are free from edited-allele bias, while the edited alleles are unaffected by LD bias. This creates a complex scenario not encountered when editing is performed in homozygous donors (**Figure 5a**).

The marginal effect of the endogenous alleles (denoted as β₀) is subject to bias from other functional variants on the same haplotype—i.e., LD bias—which is difficult to incorporate into the model. Moreover, experimentally determining the mixture ratio of endogenous and edited alleles is also challenging. However, in a special case where β₀ = β₁—indicating that the target variant is the sole functional variant within the haplotype—we can treat endogenous and edited alleles equivalently and need not distinguish between them. Under this assumption, the observed effect of the endogenous ALT allele is β₁ in the pre-editing condition; β₁ + β₂ in the ALT-to-REF editing condition; and β₁ − β₂ in the REF-to-ALT editing condition (**Figure 5a**). The three conditions can be jointly modeled to estimate both β₁ (the true caQTL effect) and β₂ (the edited-allele bias).

Notably, because this modeling approach assumes that the target variant is the sole functional variant within the haplotype prior to model fitting (i.e., we approximate the effect size of other functional variants on the same haplotype as zero), it is not suitable for achieving fine-mapping; rather, fine-mapping must already have been accomplished through other sources before applying this model. In this manuscript, we highlight several scenarios where this assumption is likely to hold: (i) the variant’s functionality has been confirmed by MPRA, (ii) the variant has already been fine-mapped in a previous study, and (iii) the variant’s causal effect has been supported by genome editing experiments using homozygous samples.

Finally, by directly estimating β₀ (the marginal effect of the endogenous allele) from pre-editing samples, we can evaluate the heterogeneity between β₀ and β₁ to assess whether our key assumption is violated. This heterogeneity test serves as a safeguard to validate the assumption that β₀ = β₁. Reassuringly, Cochran’s Q test showed no significant heterogeneity (P > 0.05) in any of the bidirectional editing experiments using heterozygous donors in this manuscript.

**Cell culture conditions**

LCL and Jurkat cells were maintained in RPMI-1640 (Wako) supplemented with 10% fetal bovine serum (FBS; Gibco, 10270-106), 100 U/mL penicillin-streptomycin (Thermo Fisher, 15140122) and one mM sodium pyruvate (Wako, 190-14881). Cells were cultured at 37 °C with 5% CO2 and tested negative for mycoplasma contamination. We activated 1x10^6^ CD4^+^ T cells with 1 ml ImmunoCult-XF T Cell Expansion Medium (STEMCELL, 10981) supplemented with 25 μL Human CD3/CD28 Activator (STEMCELL, 10971). B cells were cultured at a maximum cell density of 1x10^6^ cells/mL using the human B Cell expansion kit (Myltenyi, 130-106-196). At the start of stimulation and 0.5, 1, 2, 3, 4, 6, 8, 12, 24, 48 and 72 hours after stimulation, 20,000 cells were stored in CryoStor CS10 (Sigma-Aldrich, C2874-100ML) for UNIChro-seq.

**sgRNA and epegRNA for prime editing**

We prepared sgRNA- or epegRNA-expression plasmid vectors for prime editing of Jurkat cell lines using the protocol described previously^1^. sgRNA-expression vectors were prepared by KLD cloning. Briefly, we conducted PCR with primers containing the target sgRNA sequence and pFYF1548 EMX1 (Addgene, #47508) as a template. The PCR product was purified with QIAquick PCR Purification Kit (QIAGEN), incubated with KLD Enzyme mix (NEB M0554S) and transformed into One Shot TOP10 Chemically Competent E. coli (Thermo Fisher). For epegRNA-expression vectors, oligonucleotides for the spacer sequence, guide RNA scaffold, and 3’ extension were annealed, and assembled with BsmBI (NEB, R0739S)-digested acceptor vector (pU6-tevopreq1-GG-acceptor; Addgene #174038) using T4 DNA ligase (NEB, M0202S) according to the manufacturer’s protocol. Assembled plasmids were transformed into One Shot TOP10 Chemically Competent E.coli (Thermo Fisher) and grown on Luria-Bertani (LB) agar (Sigma, L2897) with 100 μg/ml ampicillin (Sjgma, A0166). All plasmid sequences were verified by Sanger sequencing (Eurofin). All vectors were purified using Plasmid Plus Midiprep kits (Qiagen).

For prime editing of primary T cells, we used synthetic sgRNAs and pegRNAs (Integrated DNA Technologies). The pegRNAs contained 2’-O-methyl modifications at the first and last three nucleotides and phosphorothioate linkages between the first three and last nucleotides. The sgRNAs contained 2’-O-methyl modifications at the first three and last nucleotides and phosphorothioate linkages between the first three and last two nucleotides. Sequences of pegRNAs, epegRNAs, gRNAs and sgRNAs used in this work are listed in **Supplementary Tables 17**.

**Primer design strategy**

For targeted sequencing of variant-containing regions, we designed nested primers based on genomic coordinates of variants and their associated open chromatin regions. For each target variant, we extracted 200 bp sequences from the GRCh38 reference genome, positioning the variant at the terminal end of the fragment. Sequences were processed in their forward or reverse complement orientation depending on the desired strand for primer design. We used the R package openPrimeR^2^ (v1.20.0) to design primers with the following parameters: melting temperature 62 ± 3°C (rescue parameters: 62 ± 5°C), primer length 18-22 bp (rescue: 15-25 bp), maximum primer pair melting temperature difference 5°C (rescue: 7.5°C), minimum self-dimerization ΔG -15 kcal/mol (rescue: -17 kcal/mol), minimum secondary structure ΔG -11 kcal/mol (rescue: -12 kcal/mol), and minimum cross-dimerization ΔG -17 kcal/mol (rescue: -19 kcal/mol). A coverage parameter (CVG=0.8) controlled design stringency, with higher values relaxing constraints to ensure primers for all targets. Our nested PCR strategy involved designing inner primers within a 50 bp window near the variant and outer primers positioned upstream, allowing a 10 bp overlap between primer pairs. We evaluated primer specificity by aligning both full-length sequences and 3'-terminal 15 bp regions to the GRCh38 reference genome and assessed off-target binding potential to select primers with optimal genomic specificity. The sequences of UNIChro-seq nested-PCR primers for each target locus are listed in **Supplementary Table 19.**

**Supplementary Figures**

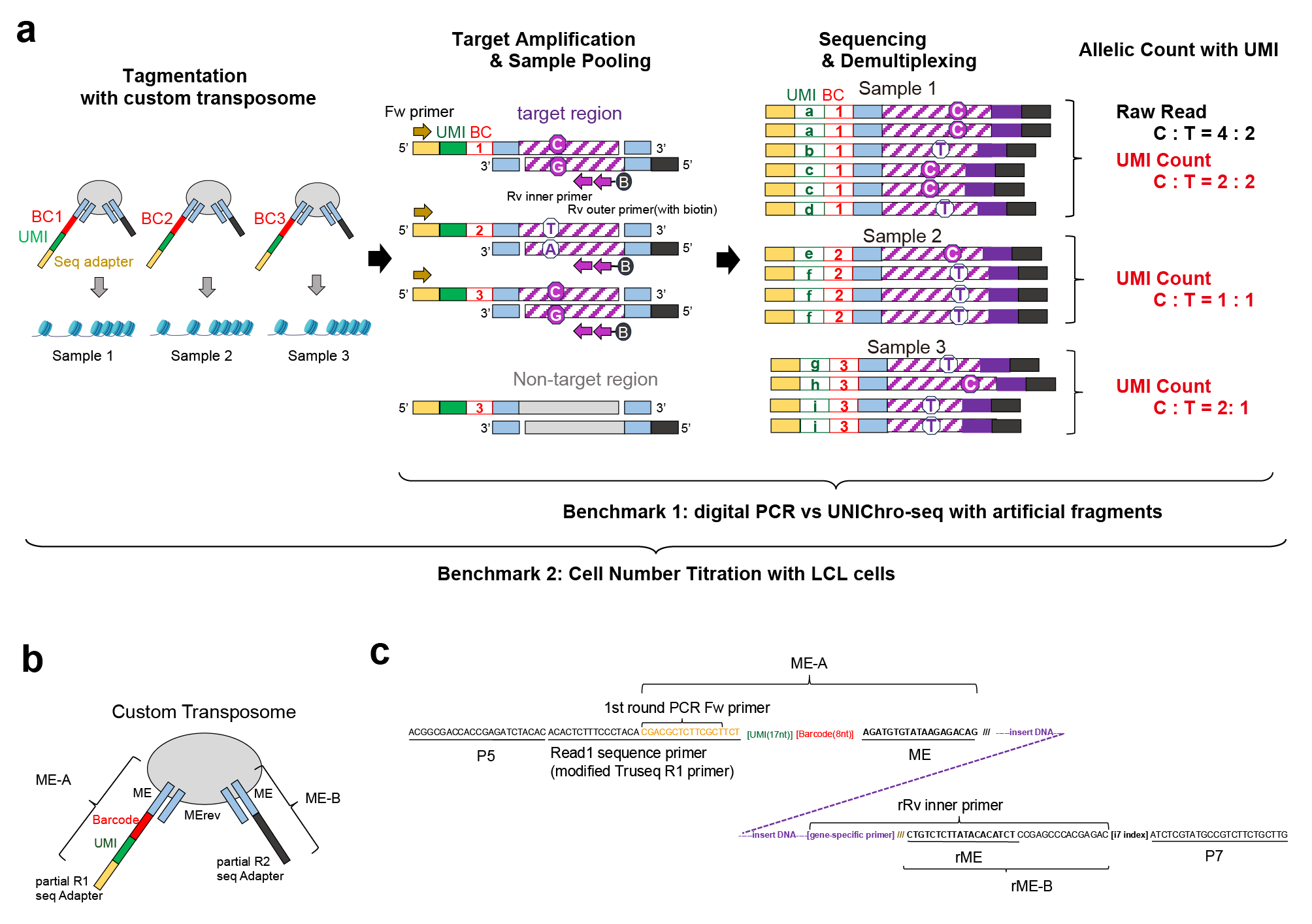

**Supplementary Figure 1: The workflow of UNIChro-seq**

**a**, The panel illustrates the experimental workflow of UNIChro-seq. Samples are digested and labeled with barcodes and UMIs, and the target regions within the tagmented fragments are amplified through three steps. The barcode enables the pooling of samples during library preparation and their subsequent demultiplexing after sequencing. The UMIs facilitate accurate assessment of the allelic ratio at the target regions by digital counting. **b**, Structure of the custom transposome used for UNIChro-seq **c**, Sequences of the UNIChro-seq library including custom transposase adapters.

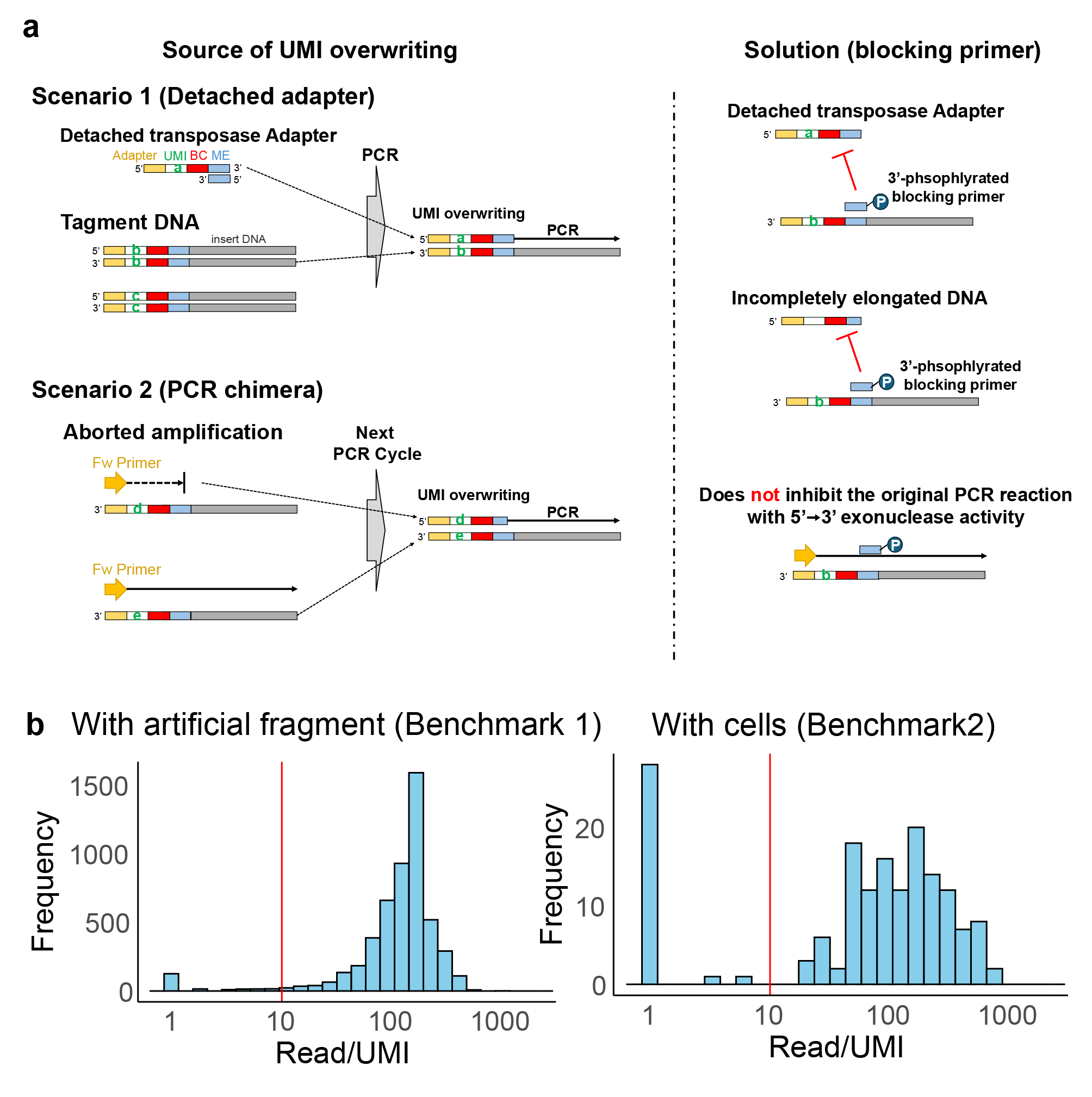

**Supplementary Figure 2: Experimental and analytical strategy to prevent unintended UMI overwriting**

**a,** The source of UMI overwriting (left) and prevention by 3’-phosphorylated blocking primer. Detached transposase adapters or partially extended primers can act as primers and overwrite UMIs (left). The 3’-phosphorylated blocking primer competitively inhibits annealing of transposase adapters and incompletely elongated primers. **b**, The number of reads of each UMI from the library with artificial fragment (benchmark 1; left) and with LCL cells (benchmark 2; right) was plotted by histogram. The red line represents the minimum cut-off used as a QC filter in this analysis.

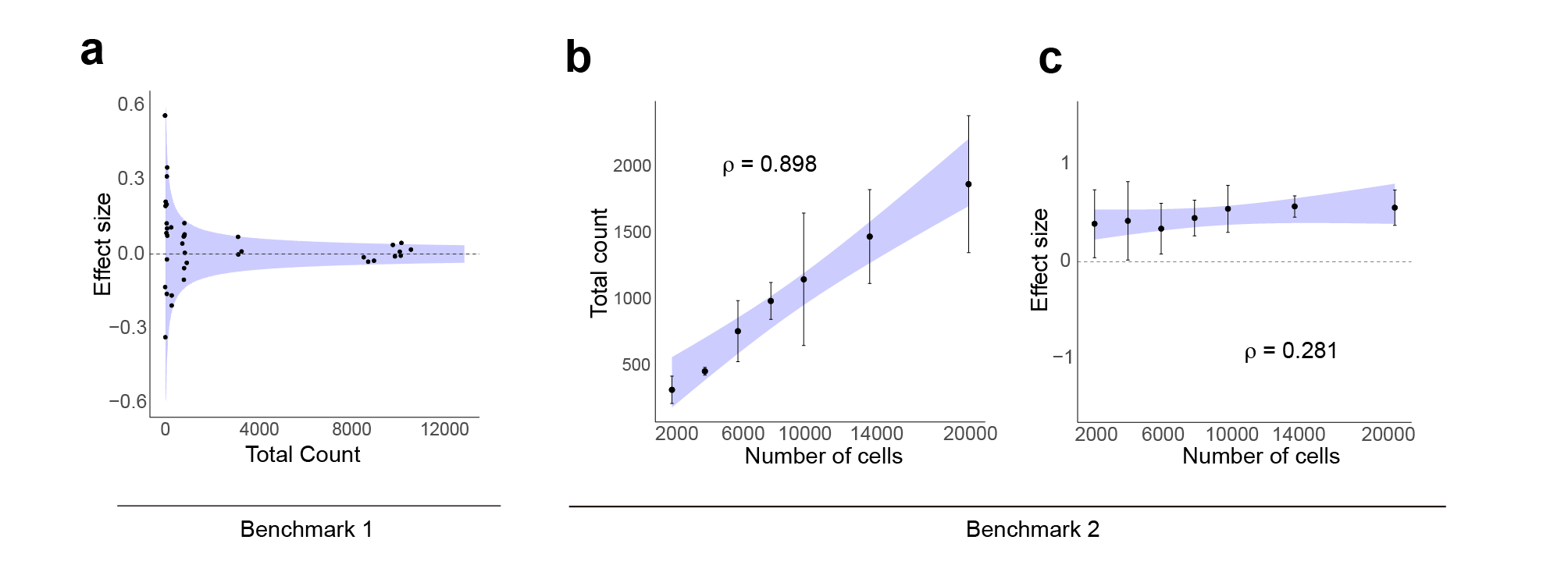

**Supplementary Figure 3. Benchmarking of UNIChro-seq**

**a**, The effect size of the C allele detected by UNIChro-seq across different copies of input artificial fragments. The blue shaded area represents the theoretical range of variation expected based on the binomial distribution. **b**, The total number of UMIs detected by UNIChro-seq across different numbers of input cells. **c**, The effect size the C allele detected by UNIChro-seq across different numbers of input cells. For panels **b** and **c,** Spearman’s ρ is provided. Error bars represent 95% confidence based on the standard error of the mean (SEM).

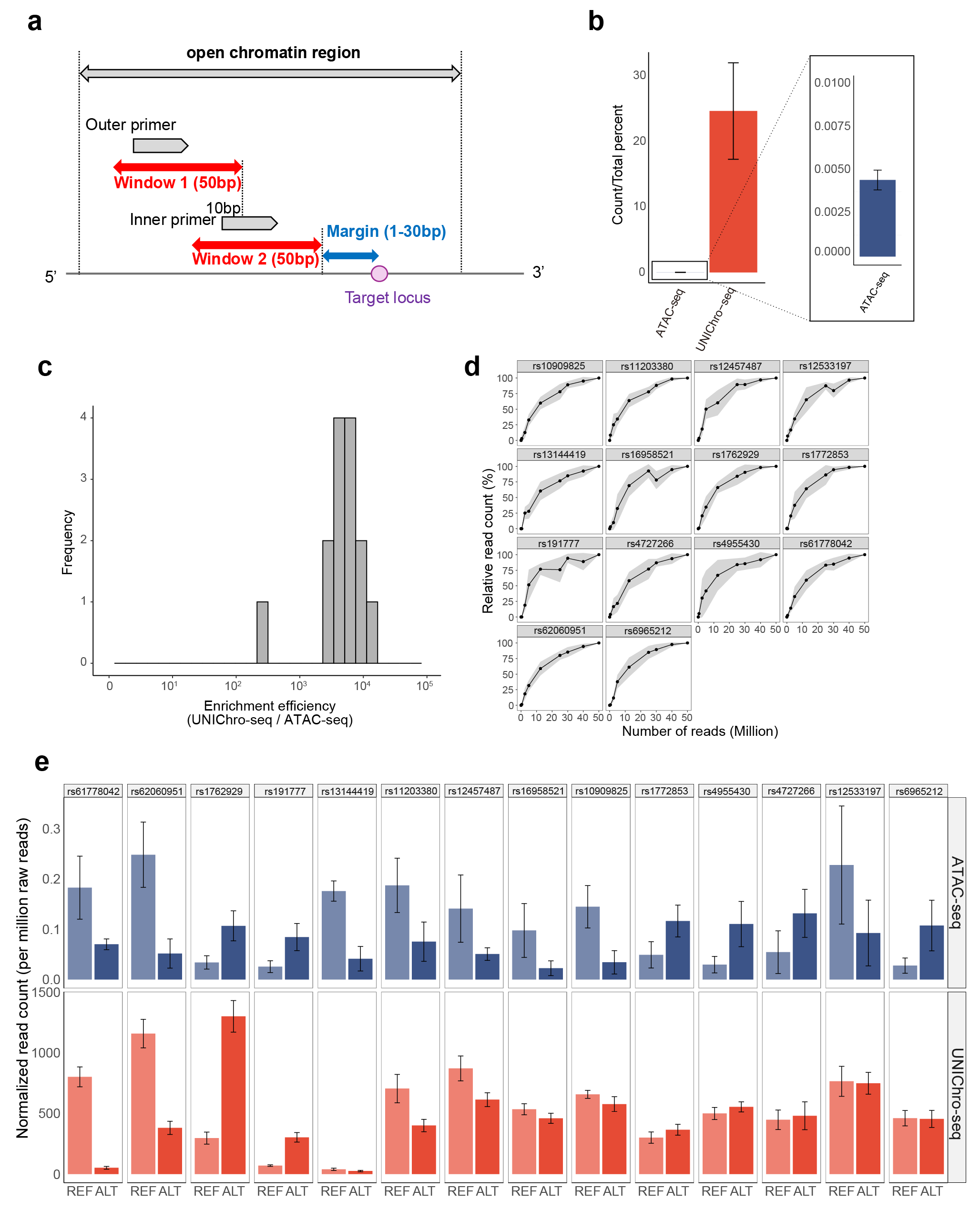

**Supplementary Figure 4. Validation of multi-target UNIChro-seq**

**a,** Primer design scheme for UNIChro-seq. We designed the inner primers within a 50-base window further out in the range of 1 to 30 bases from the target SNP, then outer primers were designed within the 50 base window outside the inner primer. Outer primers were allowed to overlap up to 10 bases with inner primers. **b**, The fraction of the reads that mapped to the 17 target regions. **c,** The enrichment efficiency of UNIChro-seq was divided by that of conventional ATAC-seq for each target variant (n=14), and the value was depicted by a histogram. **d**, To visualize all loci (n = 14) on the same scale, read counts at each sequencing depth were expressed as a percentage relative to those at 50 million reads. Each panel represents a different target locus. The solid line shows the mean normalized count, and the gray shaded area represents the 95% confidence interval calculated from the standard error of the mean (SEM). **e**, De-duplicated read count normalized to one million raw reads carrying reference and alternative allele of each target locus by conventional ATAC-seq (n = 8 replicates) and UNIChro-seq (n = 3 replicates). The error bars represent 95% confidence based on the SEM.

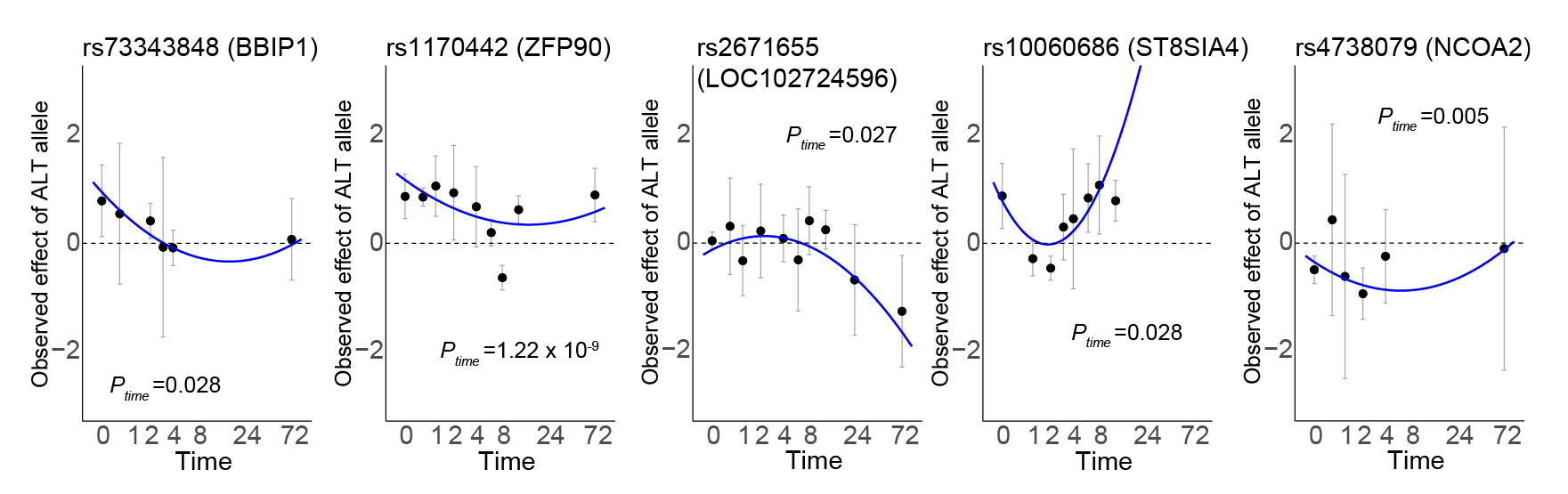

**Supplementary Figure 5. Dynamic caQTL effect of SLE GWAS variants.**

The dynamic caQTL effect of SLE risk variants with nominal P < 0.05 that passed QC. Error bars indicate 95% confidence interval from mixed-effects logistic regression fitted independently at each time point. P_time indicates the P-value for the linear time-dependent caQTL effect from model 2 (See Method)..

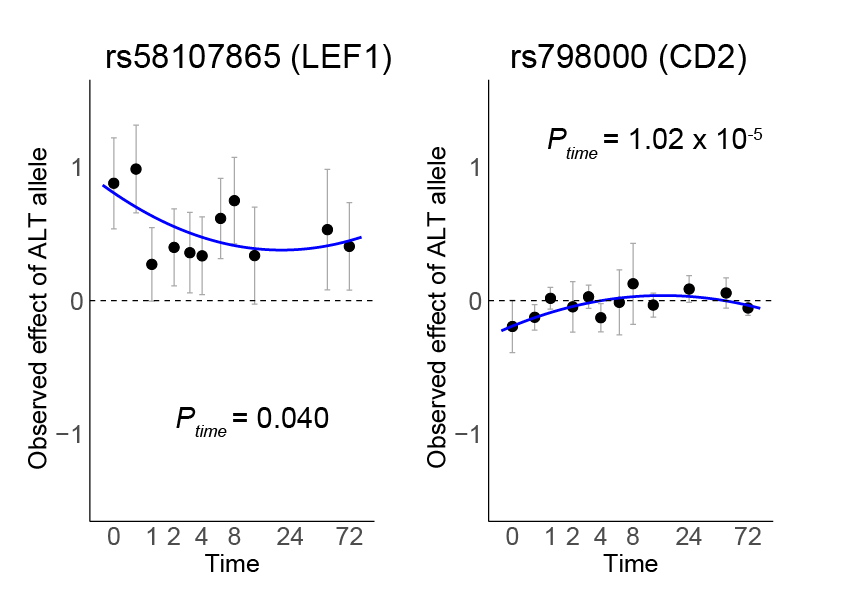

**Supplementary Figure 6. Dynamic caQTL effect of RA GWAS variants.**

The dynamic caQTL effect of RA risk variants with nominal P < 0.05 that passed QC. Error bars indicate 95% confidence interval from mixed-effects logistic regression fitted independently at each time point. P_time indicates the P-value for the linear time-dependent caQTL effect from model 2 (See Method).

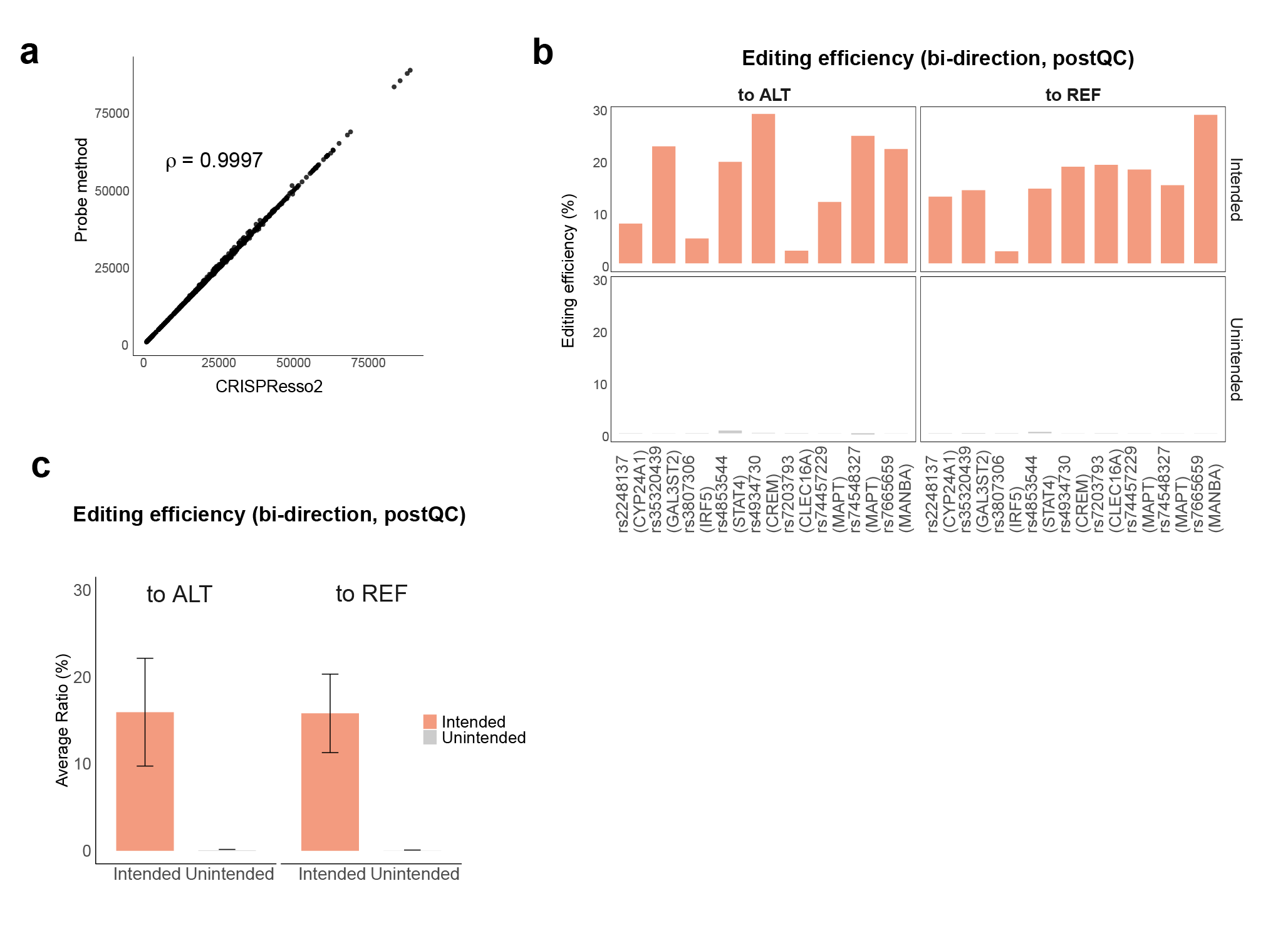

**Supplementary Figure 7. Uni-directional and bi-directional editing for detecting true caQTL effect of GWAS variants.**

**a,** Comparison of the number of reads detected as intended edits with CRISPResso2 and our in-silico probe method. The Spearman’s ρ is provided. **b**, Editing status of bi-directional editing across the editing direction in Jurkat cells after QC. The mean of eight biological replicates is provided for each variant (n=9). **c**, The mean bi-directional editing efficiency for nine variants.

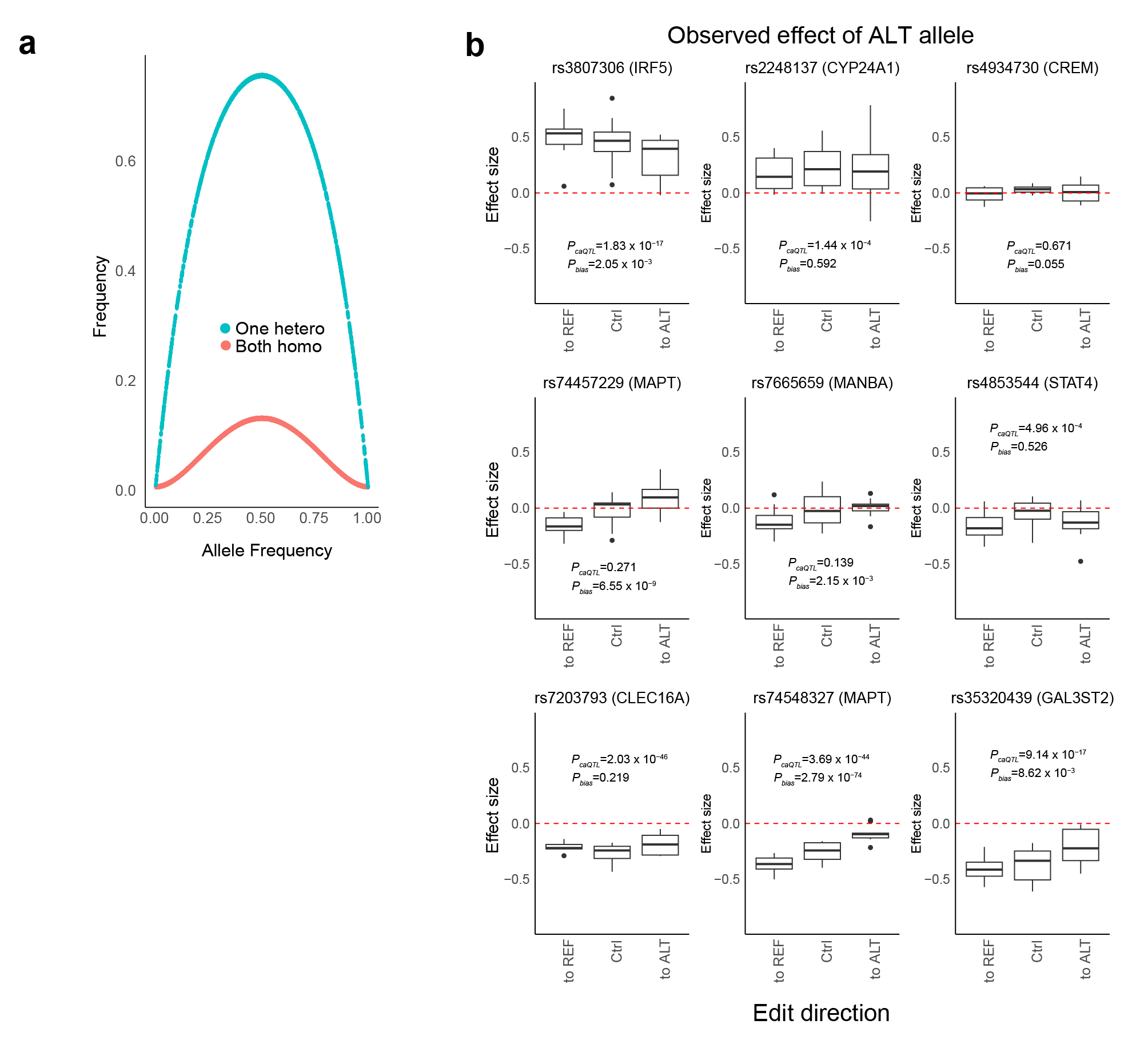

**Supplementary Figure 8. CaQTL effect detected by bi-directional editing of GWAS variants**

**a,** Assuming we recruit two donors, we calculated the frequency of having at least one heterozygous donor and that of having donors with two homozygous genotypes (REF/REF and ALT/ALT). **b**, Effect size estimate of target loci across editing direction. *P*-value for caQTL (*P_caQTL_*) and edited-allele bias (*P_bias_*) estimated by a joint model is provided.

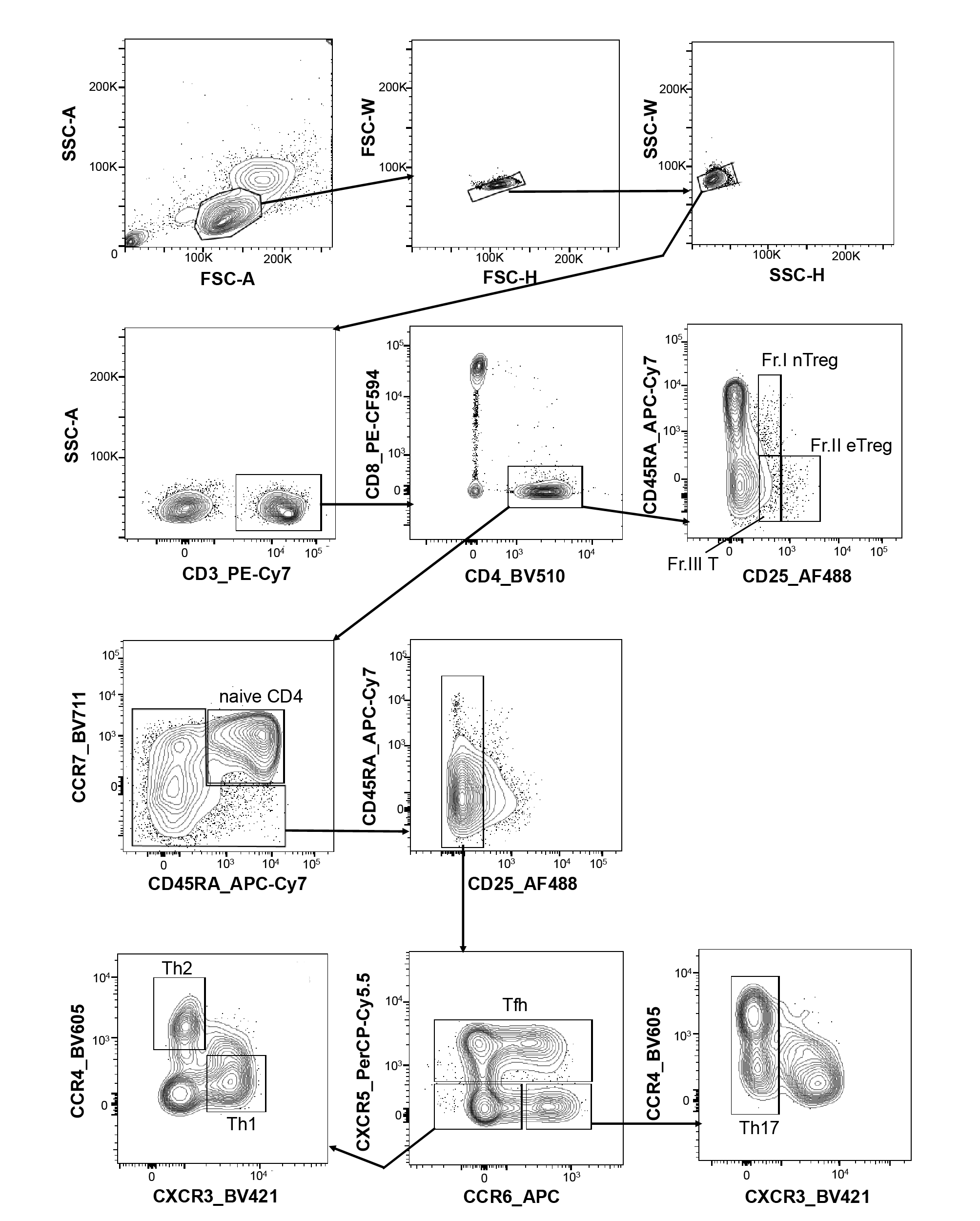

**Supplementary Figure 9. Gating strategy of CD4+ T cell subset sorting**

Representative FACS plots describing gating strategies for CD4+ T cell subset sorting. Th1, T helper 1 cells; Th2, T helper 2 cells; Th17, T helper 17 cells; Tfh, T follicular helper cells; Fr.I nTreg, Fraction I naïve regulatory T cells; Fr.II nTreg, Fraction II effector regulatory T cells; Fr.III T, Fraction III non-regulatory T cells.

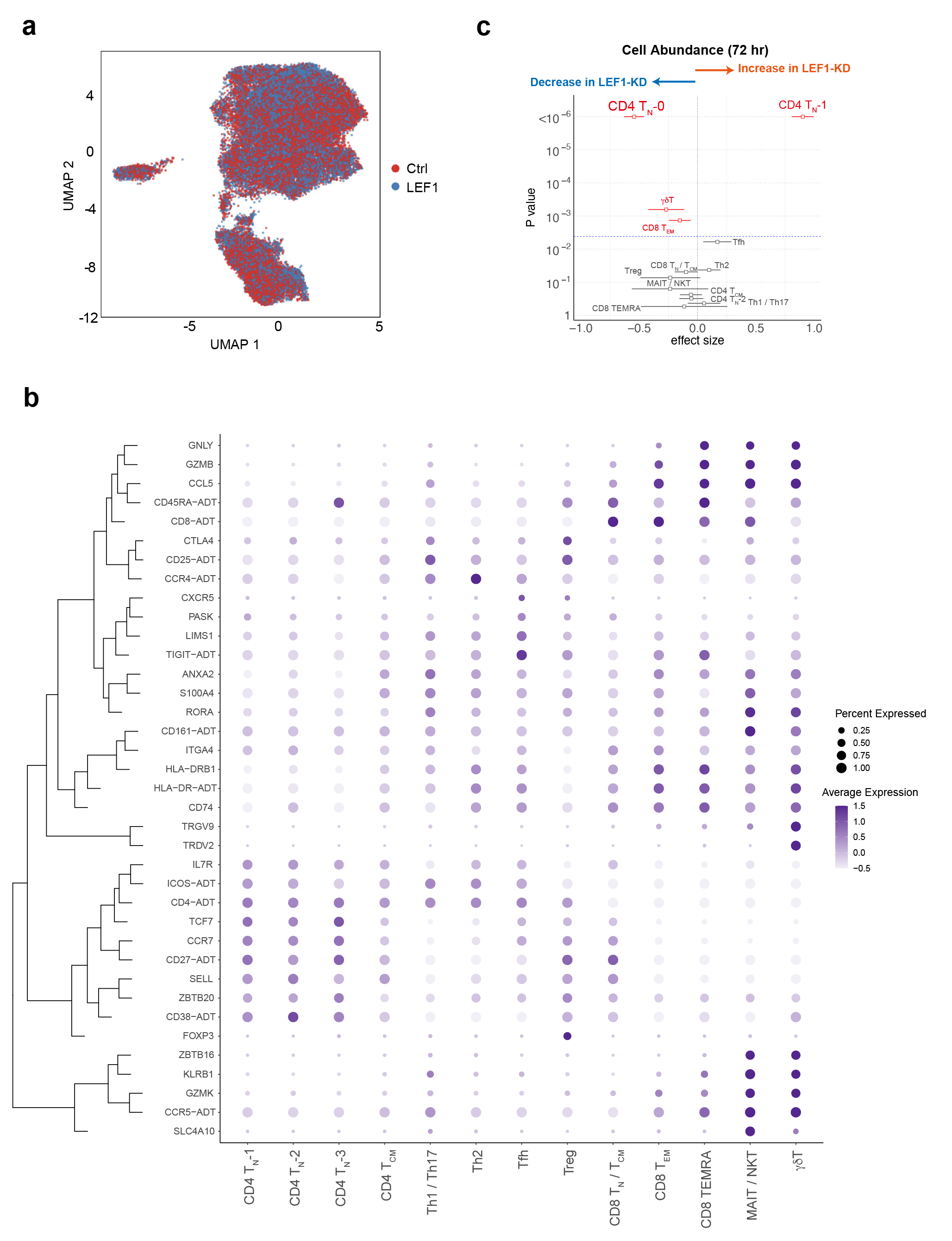

**Supplementary Figure 10. Single cell multi-omics analysis with LEF1 knockdown T cells**

**a,** A UMAP embedding of CITE-seq data for *LEF1*-knockdown and control T cells. Colors represent experimental conditions. **b,** The shift in the cell type abundance by *LEF1* knockdown at 72h post-transfection estimated by MASC^3^. Cell clusters with a significant shift are highlighted in red. The dashed line indicates the significance threshold (*P* = 0.05/13). **c**, A dot plot of marker genes across all cell clusters. The shade of purple indicates the average expression in each cell cluster and the dot size represents the proportion of cells expressing the gene.

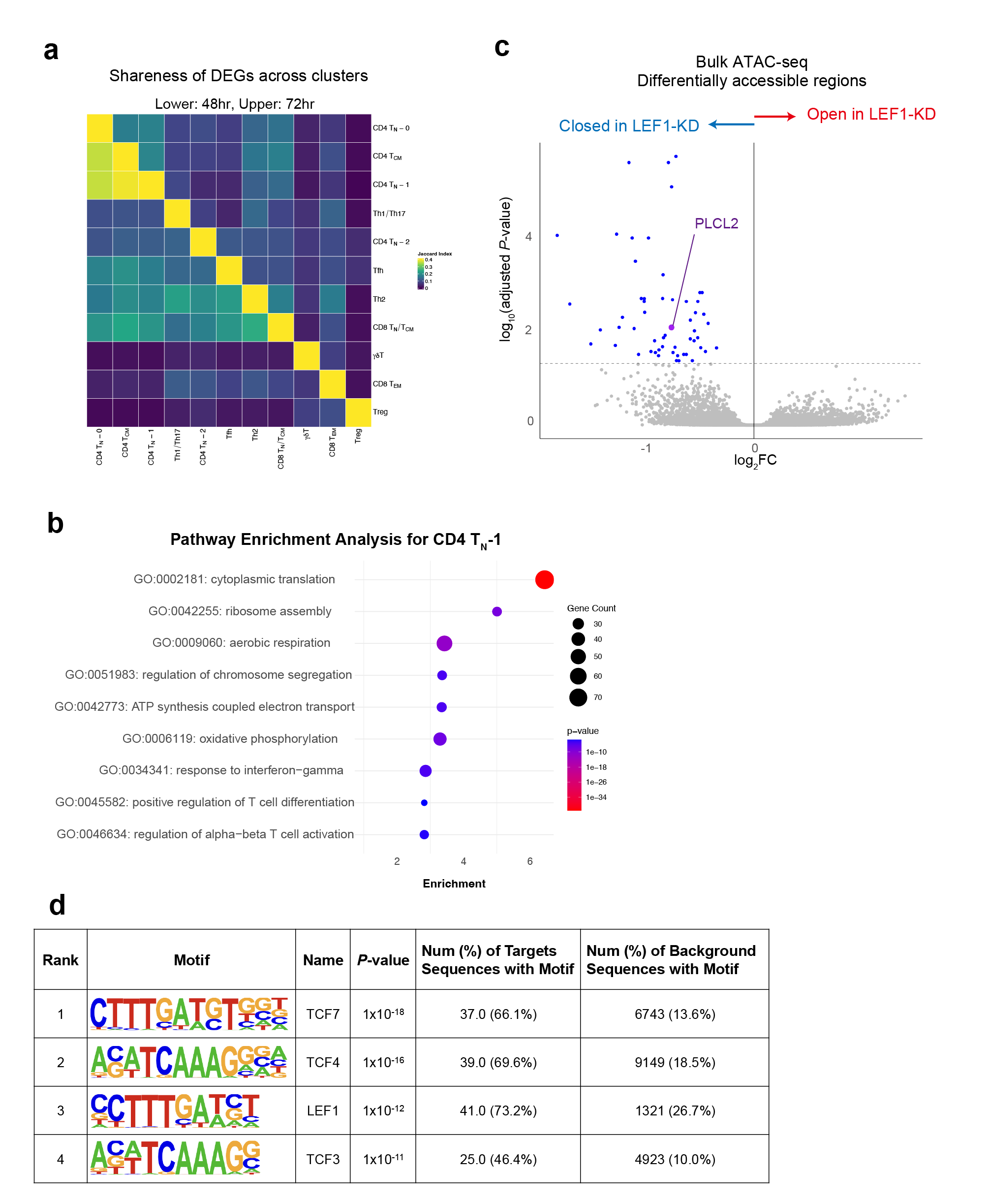

**Supplementary Figure 11. The impact of LEF1 knockdown in T cells**

**a**, Pairwise comparison of differentially expressed genes (FDR < 0.05) shared between all cell clusters in this study. **b**, Differentially accessible regions detected by conventional ATAC-seq. Blue dot indicates the region with significance (FDR < 0.05). **c**, Pathway enrichment of the DEGs in CD4 T_N_-1 cluster by LEF1 knockdown. Dot represents the number of DEGs detected in each pathway and the color shows *P*-value in Fisher’s exact test. d. Transcriptional factor motif enrichment analysis of differentially accessible regions by LEF1-silencing in CD4+ T cells.
